# Modeling the Cost-Effectiveness of the COVID-19 mRNA-1273 vaccine in the United States

**DOI:** 10.64898/2025.12.12.25342164

**Authors:** Kelly Fust, Ekkehard Beck, Michele Kohli, Shannon Cartier, Nicolas Van de Velde, Milton Weinstein, Keya Joshi

## Abstract

**Objective:** The main objective was to estimate the potential public health impact and cost-effectiveness of an annual dose of mRNA-1273 (2025/2026 formula) in the United States (US) for the 2025-2026 season compared with no vaccination in the mRNA-1273 licensed population (6 months-64 years with underlying medical conditions and all ≥65 years). mRNA-1273 was also compared to BNT162b2 in high-risk adults ages 18-64 years and all ≥65 years.

**Methods:** Analyses were conducted using a previously developed static decision-analytic model (1-year horizon) from the societal cost perspective. Vaccine effectiveness (VE) against infection and hospitalization for mRNA-1273 versus no vaccination was based on a 2024-2025 real world effectiveness study. VE estimates for mRNA-1273 versus BNT162b2 were based on systematic literature reviews and meta-analyses. Cost-effectiveness was assessed in terms of incremental cost per quality-adjusted life-year (QALY) gained and the benefit cost ratio (BCR) in the licensed target population as well as age-specific subgroups. Sensitivity and scenario analyses were performed.

**Results:** The incremental cost per QALY gained for mRNA-1273 compared to no vaccine was $23,265. For every 1 USD of mRNA-1273 vaccine related costs, there is a return of 1.91-7.90 dollars in societal perspective cost savings and monetized health benefit gained. In the subgroup of high-risk individuals 6 months-4 years, mRNA-1273 was associated with lower costs and improved health outcomes, resulting in mRNA-1273 dominating no vaccine. Study results are sensitive to COVID-19 incidence, percentage hospitalized, post-discharge mortality, and VE assumptions.

Compared to BNT162b2, given improved clinical outcomes, combined with a lower vaccine unit cost, mRNA-1273 was shown to dominate BNT162b2.

**Conclusions:** mRNA-1273, the only licensed vaccine for those <5 years of age at high risk of severe COVID-19 related outcomes, could substantially reduce the clinical and economic burden of COVID-19 among US high-risk populations and older adults. These benefits were observed both in comparison to no vaccination and the BNT162b2 vaccine.

## Introduction

The Centers for Disease Control and Prevention (CDC) in the United States (US) has stated that individuals aged 6 months and above may receive at least one COVID-19 vaccine in 2025-2026 using shared clinical decision making.^1^ The only Food and Drug Administration (FDA) approved vaccine for all age groups is mRNA-1273 (Spikevax, Moderna, Inc.), which is indicated for use in those 6 months through 64 years of age with at least one underlying condition that puts them at high risk for severe outcomes from COVID-19 and those 65 years of age and older.^2^ The 2025-2026 formula of this vaccine targets the JN.1-lineage LP.8.1 strain.^3^ The main objective of this manuscript was to estimate the potential public health impact and cost-effectiveness of mRNA-1273 (2025-2026 formula) in the US for the 2025-2026 season compared with no vaccination. A secondary objective was to compare mRNA-1273 to BNT162b2 (Comirnaty, BioNTech Manufacturing GmbH / Pfizer Inc.), which has a similar indication as mRNA-1273, except that it is limited to those 5 years and older.^4^

## Methods

### Overview

A previously described Markov model with monthly cycles, described in detail by Fust et al. (2025)^5,6^, populated for the US setting, was used to project the potential clinical outcomes (number of symptomatic COVID-19 infections, hospitalizations, deaths and cases of long COVID) and societal perspective economic outcomes (quality-adjusted life-year (QALY) losses and costs) expected in the mRNA-1273 indicated population. Analyses were performed over a one-year time horizon (September 2025-August 2026) with a single dose of mRNA-1273 (annual vaccination) compared to no vaccination.^7–9^ The number of antibiotic prescriptions in the outpatient setting avoided through mRNA-1273 vaccination was estimated.^6^ Incremental cost-effectiveness ratios (ICERs) were calculated as the incremental cost per QALY gained. Results are reported for the overall target population, and the high-risk 6 months-4 years, high-risk 5 years to 64 years, and all ≥65 years subgroups. As the CDC guidelines state that those ≥65 years may receive a second dose a minimum of two months after the first dose (semi-annual vaccination),^10^ a scenario analysis where those ≥65 years receive 2 doses was conducted.

Vaccination with mRNA-1273 was compared to vaccination with BNT162b2 in the population of high-risk adults ages 18-64 and all adults ages ≥65 years; this analysis was limited to those ≥18 years of age because BNT162b2 is not licensed for those ages 6 months-4 years at high-risk, and due to a lack of comparative data between mRNA-1273 and BNT162b2 in the pediatric population and lack of 1273-specific VE data for those <18 years of age.^11^ Benefit cost ratios (BCRs) were calculated as the benefit of vaccination (cost savings from averting infections and monetized health gains due to averted COVID-19 outcomes) divided by the costs of vaccination (costs and monetized health lost due to vaccine adverse events (AEs)). Health was monetized using various assumptions: a QALY gained was valued at $100,000 and $150,000 (standard willingness-to-pay thresholds for a QALY in the US)^12^; value per statistical life-year (VSLY) $604,000^13^ and value per QALY (VQALY) $717,000.^13^ A completed CHEERS checklist can be found in the Technical Appendix.

For the analyses comparing mRNA-1273 and BNT-162b2, a range of relative VE (rVE) values were used to calculate the ranges used for sensitivity analyses as shown in Table 1; scenario analyses examining COVID-19 incidence and vaccine coverage were also performed.

**Table 1.** Initial COVID-19 Vaccine Effectiveness.

| Age Group (Years) | Symptomatic Infection |  |
| --- | --- | --- |
|  | 2024-2025 mRNA-1273 VE (95% CI)* | BNT-162b2VE (Range)** |
| 6 mos – 4 High-Risk | 84.4%<br>(22.4%, 100%) | -- |
| 5 -17 High-Risk | 62.4%<br>(38.4%; 77.4%) | -- |
| 18 – 64 High-Risk | 48.2%<br>(43.3%; 52.7%) | 41.8%<br>(35.3% - 47.7%) |
| ≥65 | 54.5%<br>(49.5%; 59.0%) | 38.5%<br>(26.6% - 48.3%) |
|  | Hospitalization<sup>†</sup> |  |
| 6 mos – 4 High-Risk | 84.4%<br>(22.4%, 100%) | -- |
| 5 -17 High-Risk | 62.4%<br>(38.4%; 77.4%) | -- |
| 18 – 64 High-Risk | 50.5%<br>(29.3%; 65.7%) | 38.2%<br>(30.3% - 45.7%) |
| ≥65 | 57.2%<br>(38.2%; 70.7%) | 38.0%<br>(19.3% - 52.0%) |

CI: Confidence interval; RR: Relative risk; VE: Vaccine effectiveness; -- Not applicable
\*For those ages 18 years and older, VE against infection and hospitalization were measured at 64 days post administration and were scaled back to 14 days post administration to estimate the initial VE in the model as described in the Technical Appendix. For those 6 months to 17 years, values were measured at 81 days in the RWE study, so the observed VE estimate was scaled back 67 days to 14 days post-administration conservatively assuming the monthly VE waning rate against hospitalization<sup>23</sup>
\*\* BNT-162b2 was calculated using the base case 1273 VE and a RR of 0.89 (18-64 years high-risk; 95% CI: 0.80 – 0.99) and 0.74 (≥65 years; 95% CI: 0.62-0.88) for infection and a RR of 0.80 (18-64 years high-risk; 95% CI: 0.71-0.91) and 0.69 (≥65 years; 95% CI: 0.53-0.89).
<sup>†</sup> If hospitalization is lower than infection VE, the incremental protection against the probability of hospitalization given infection is set to 0.

### Target Population Size

A total of 239,582,213 people, including persons ≥65 years in addition to eligible 6 months-64-year olds, were modeled based on the US population^14^ and FDA licensure for mRNA-1273.^15^ For ages 6 months – 64 years, the cohort size eligible for vaccination is limited to high-risk individuals, estimated from Forrest et al. (2025)^16^ (6 months – 17 years) and Panagiotakopoulos (2025)^17^ (18-64 years). (See Appendix Section 3)

### Model Structure and Inputs

The model structure was previously described in publications by Fust et al.^6^ Briefly, individuals are subject to risk of infection based on estimated US age-specific incidence of SARS-CoV-2 (average of 2023-2024 and 2024-2025 seasons) on a monthly basis. If those individuals acquire infection, a decision tree is used to determine which COVID-19 outcomes they develop following infection. Each month, individuals may receive a COVID-19 vaccine according to age-specific coverage rates. Vaccination reduces the risk of COVID-19 related infection and hospitalization following infection; sensitivity analyses considered a multiplier effect to reflect prevention of transmission as an approximation of indirect benefit.

The derivation of model probabilities, costs, and utility inputs were previously described.^6^ Any changes from this previous analysis, including inputs for the pediatric population (ages 6 months to 17 years), are described in this manuscript. The approach to estimating monthly incidence of symptomatic infection in the target population (assuming no 2025-2026 vaccination) and monthly vaccine coverage are consistent with Fust et al. (Technical Appendix Sec. 1). The remaining model probabilities, including the probability of vaccine-related adverse events (AEs), are described in the Technical Appendix. COVID-19 related QALY losses and AEs were assumed to be the same for the general population and high-risk patients and are consistent with Fust et al.^6^ Cost estimates are presented in 2025 US dollars and are summarized in the Appendix. Non-health care costs for COVID-19 patients included lost productivity from time missed from work due to receiving a COVID-19 vaccine and AEs, COVID-19 infection and long COVID, and infection-induced myocarditis. Caregiver productivity loss was also included to account for time loss from work to care for patients with COVID-19. It was assumed that patients <18 years would require caregiver time for vaccination, vaccination-related AEs, and during the acute infection period, and that those ≥18 years of age would require a caregiver for long COVID only. *Vaccine Effectiveness*

All vaccine effectiveness (VE) inputs for the first month of the time horizon (initial VEs) are shown in Table 1. mRNA-1273 VEs were based on a large, US nationwide 2024-2025 real world effectiveness (RWE) study providing interim VE estimates of mRNA-1273.712 targeting KP.2.^18^ Estimates for high-risk populations were used for those aged 18-64 years. For those <18 years, the 2024-2025 VEs for mRNA-1273 against infection and hospitalization were assumed to be the same and were based on MacNeil’s estimates of the RWE of 2024-2025 COVID-19 vaccines against emergency department and urgent care visits in children, as these were the only available US VE estimates applicable to mRNA-1273 for the pediatric population.^19^

For scenario analyses comparing mRNA-1273 and BNT-162b2, data from two systematic literature reviews (SLR) and meta-analyses were used. Wang et al. (2025) compared mRNA-1273 and BNT-162b2 in adults with underlying medical conditions and found those aged ≥18 years vaccinated with mRNA-1273 had significantly lower risks of SARS-CoV-2 infection and hospitalization.^20^ Another SLR and meta-analysis in older adults, by Kavikondala et al. (2024),^21^ found similar results, with those ≥50 years receiving mRNA-1273 having reduced risk of SARS-CoV-2 infection and hospitalizations. For sensitivity analyses, the VE of mRNA-1273 was held constant while the RRs were varied using the 95% confidence intervals to calculate low and high values of VE for BNT-162b2.

In the model, VE declines linearly on a monthly basis. Consistent with Fust et al.^6^, the monthly VE waning of mRNA vaccines against infection was estimated to be 4.75% (95% CI 3.05%; 6.75%) based on a meta-analysis by Higdon et al. (2022).^22^ The base case rate of waning of VE against hospitalization of 2.46% per month was estimated from Andersson et al., (2024)^23^, while ranges for sensitivity analyses came from Higdon et al against hospitalization (1.37%) and an alternate study of the mRNA-1273 XBB.1.5 vaccine (3.87%).^24^

### Vaccine Unit Costs

Unit costs of the vaccines, by age group, were estimated based on 2025/2026 season list prices.^25^ For mRNA-1273, unit prices for ages 6 months–11 years and 12-17 years were $78.23 and $83.76 for the CDC cost per dose and $129.00 and $141.80 for the private sector, respectively; a 50%/50% split between the CDC and private sector costs was used in model analyses.^25,26^ The resulting unit prices used in model analyses were $103.62 for mRNA-1273 in the 6 months–4 year age group and a weighted average (based on the US population) of $108.04 for the 5-17 year age group. For those ages ≥18 years, a unit cost of $141.80 was used for mRNA-1273.^25^ For BNT162b2, a unit cost of $147.69 for those ages ≥18 years was used.^25^ An administration fee of $20.05 was included in base case.^27^ A sensitivity analysis considered a vaccine administration fee of $40 based on the Centers for Medicare & Medicaid Services (CMS) national payment allowance in 2024-2025 for COVID-19 vaccine administration.^28^

### Sensitivity and Scenario Analyses

Deterministic sensitivity analyses (DSAs) were performed to assess the impact of key variables on both clinical and economic outcomes. All values except for VE (presented above) are shown in the Technical Appendix. Where available, estimates were varied according to 95% confidence intervals (CIs) or reported ranges. For all other DSAs, an alternative data source was used or parameters were varied by ±10% or ±25% of their base-case value.

The same scenario analyses described in Fust et al.^6^ were repeated to test the impact of specific changes to VE inputs, to estimate the potential impact of changes in transmission (i.e. indirect effect) following vaccination, to increase vaccine coverage rates in September by 5% or 10% for all ages and for those ≥18 years only, and to change the percentage of patients seeking no formal care to be equal to CDC influenza rates from the 2023-2024 season. In addition, for high-risk individuals, scenarios were performed assuming that the probability of receiving outpatient care equivalent to the general population, the RR of inpatient mortality is 1.62 based on cardiovascular disease^29^, and the percentage hospitalized was based on the RR derived from Wilson et al. (2025) (RR=1.45). Scenario analysis considering the healthcare perspective, as well as the societal perspective adding non-market productivity losses associated with premature mortality, excluding all productivity losses (market and non-market) associated with premature mortality, excluding caregiver productivity losses, and considering opportunity costs as described in Fust et al.^6^ were also conducted. A scenario analysis was also performed for the 6 months – 4 year age group only assuming double the unit price of mRNA-1273 (as a proxy for the receipt of two priming doses).

## Results

### Base case analysis (compared to no vaccine)

As shown in Table 2, use of an annual dose of mRNA-1273 is estimated to reduce the number of symptomatic infections, hospitalizations, and deaths in the target population by 2.0 million, 101,739, and 13,285 compared to no vaccine, respectively. In the base case, the numbers needed to vaccinate (NNV) to avert one COVID-19 symptomatic infection, outpatient episode, hospitalization, death and long COVID were estimated at 31, 109, 616, 4,717, and 1,158, respectively. In the subgroup of high-risk children ages 6 months to 4 years, mRNA-1273 prevented 131,850 symptomatic infections (2.9% reduction compared to no vaccine), 553 hospitalizations (3.4% reduction), and 46 deaths (3.4% reduction). Given the hospitalizations avoided, mRNA-1273 compared to no vaccine was estimated to prevent 187,200 hospital bed days in the target population during crowded conditions (additional details provided in Fust et al.^6^). The number of antibiotic prescriptions avoided in the outpatient setting with use of mRNA-1273 compared to no vaccine is 145,020.

**Table 2:** Health outcomes and health outcomes averted.

| Strategy | Health outcomes* |  |  |  |  |
| --- | --- | --- | --- | --- | --- |
|  | Cases | Outpatient | Long COVID | Hosp. | Deaths |
| **Base-Case: Annual dosing for 6 months-64 years High-Risk, 65+ All** |  |  |  |  |  |
| No vaccine | 34,939,927 | 6,564,159 | 577,355 | 621,578 | 78,531 |
| mRNA-1273 | 32,947,076 | 5,991,016 | 523,214 | 519,839 | 65,246 |
| Outcomes Averted by mRNA-1273 (%) | 1,992,850 (5.7%) | 573,144 (8.7%) | 54,141 (9.4%) | 101,739 (16.4%) | 13,285 (16.9%) |
| **Base-Case for the 6 months-4 years High-Risk Subgroup: Annual dosing** |  |  |  |  |  |
| No vaccine | 4,490,489 | 238,793 | Not applicable<sup>†</sup> | 16,420 | 1,353 |
| mRNA-1273 | 4,358,639 | 231,853 | Not applicable<sup>†</sup> | 15,867 | 1,308 |
| Outcomes Averted by mRNA-1273 (%) | 131,850 (2.9%) | 6,940 (2.9%) | Not applicable<sup>†</sup> | 553 (3.4%) | 46 (3.4%) |
| **Base-Case for the 5 years – 64 years High-Risk Subgroup: Annual dosing** |  |  |  |  |  |
| No vaccine | 24,801,288 | 3,320,432 | 302,181 | 136,667 | 15,002 |
| mRNA-1273 | 23,757,132 | 3,165,019 | 287,582 | 127,024 | 13,911 |
| Outcomes Averted by mRNA-1273 (%) | 1,044,156 (4.2%) | 155,413 (4.7%) | 14,599 (4.8%) | 9,643 (7.1%) | 1,090 (7.3%) |
| **Base-Case for the 65+ years (All) Subgroup: Annual dosing** |  |  |  |  |  |
| No vaccine | 5,648,149 | 3,004,934 | 275,174 | 468,490 | 62,176 |
| mRNA-1273 | 4,831,305 | 2,594,144 | 235,633 | 376,948 | 50,027 |
| Outcomes Averted by mRNA-1273 (%) | 816,844 (14.5%) | 410,790 (13.7%) | 39,541 (14.4%) | 91,542 (19.5%) | 12,149 (19.5%) |
| **Scenario Analysis: Annual dosing for 6 months-64 years High-Risk, Semi-annual dosing for 65+ All** |  |  |  |  |  |
| No vaccine | 34,939,927 | 6,564,159 | 577,355 | 621,578 | 78,531 |
| mRNA-1273 | 32,909,139 | 5,969,318 | 521,350 | 518,207 | 65,030 |
| Outcomes Averted by mRNA-1273 (%) | 2,030,788 (5.8%) | 594,842 (9.1%) | 56,005 (9.7%) | 103,371 (16.6%) | 13,501 (17.2%) |

Hosp.: Hospitalizations
\*Due to rounding, there may be differences in the last digit of outcomes averted
<sup>†</sup>Long COVID is assumed to apply to patients ≥18 years only

Considering economic outcomes, annual dosing with mRNA-1273 in the target population (base-case analysis) is expected to lead to a gain of 129,712 QALYs and a $3 billion increase in total costs, giving an incremental cost per QALY gained of $23,265 for mRNA-1273 compared to no vaccine (Table 3). In the subgroup of high-risk children aged 6 months to 4 years, mRNA-1273 is associated with 2,866 fewer QALYs lost and $61 million in cost savings compared to no vaccine, resulting in mRNA-1273 dominating no vaccine in this subgroup, while additional investment was required for the other age sub-group (Table 3). After doubling the vaccine cost for those 6 months-4 years, mRNA-1273 still dominated no vaccine (i.e., no change from the base-case) in this subgroup (Table 4). In the scenario analysis examining a second dose for those aged ≥65 years, the ICER for mRNA-1273 was estimated to be $28,643 per QALY gained relative to no vaccine. Vaccinating with mRNA-1273 generates BCRs of 1.91 to 7.90 USD, depending on how the health benefit was monetized. In the 6 months – 4 years high-risk subgroup specifically, the BCRs ranged from 6.03 to 30.07. (See Supplement Table 36 for BCRs.)

**Table 3:**
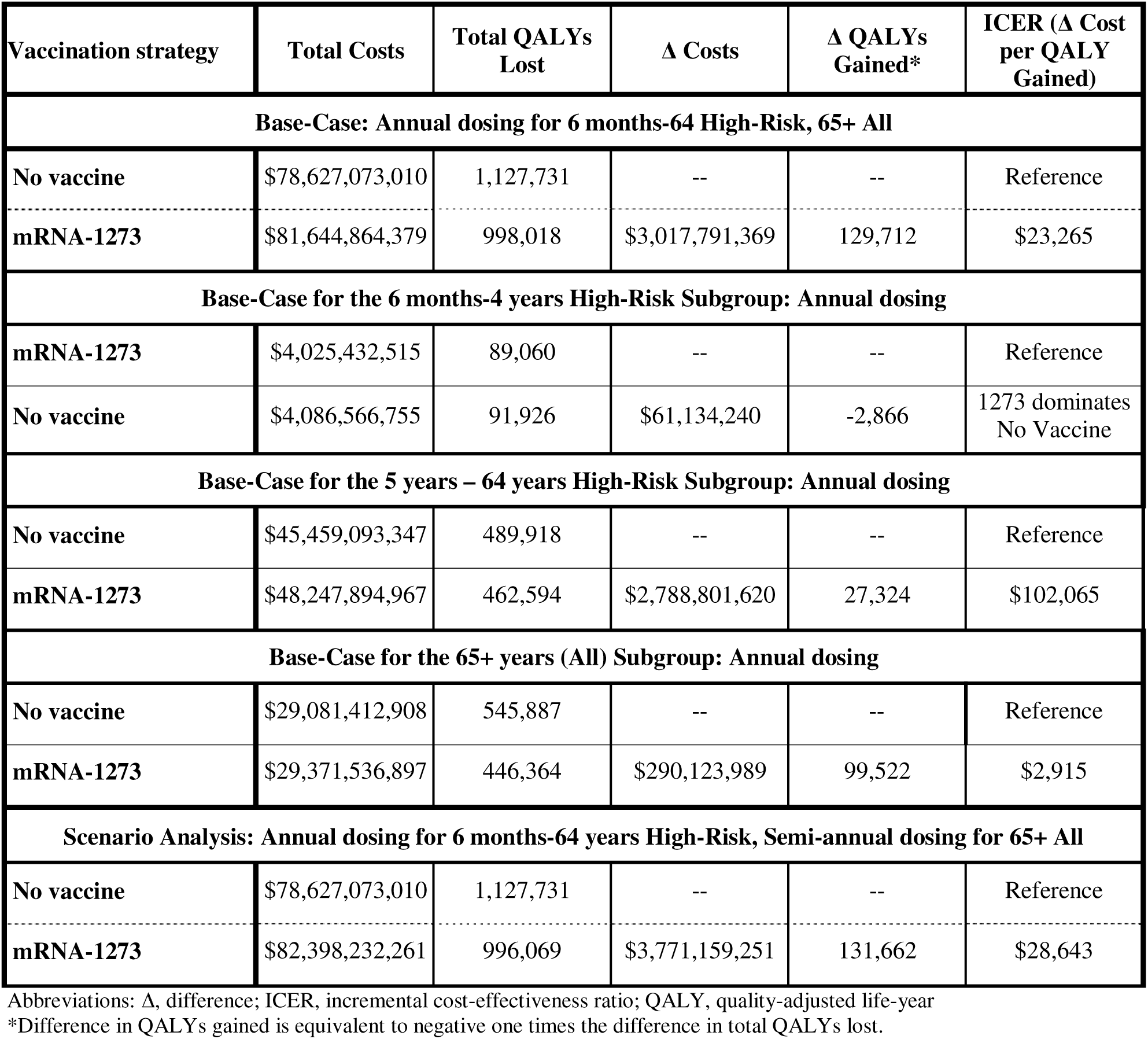
Base-case cost-effectiveness results.

| Vaccination strategy | Total Costs | Total QALYs Lost | Δ Costs | Δ QALYs Gained* | ICER (Δ Cost per QALY Gained) |
| --- | --- | --- | --- | --- | --- |
| **Base-Case: Annual dosing for 6 months-64 High-Risk, 65+ All** |  |  |  |  |  |
| No vaccine | \$78,627,073,010 | 1,127,731 | -- | -- | Reference |
| mRNA-1273 | \$81,644,864,379 | 998,018 | \$3,017,791,369 | 129,712 | \$23,265 |
| **Base-Case for the 6 months-4 years High-Risk Subgroup: Annual dosing** |  |  |  |  |  |
| mRNA-1273 | \$4,025,432,515 | 89,060 | -- | -- | Reference |
| No vaccine | \$4,086,566,755 | 91,926 | \$61,134,240 | -2,866 | 1273 dominates No Vaccine |
| **Base-Case for the 5 years – 64 years High-Risk Subgroup: Annual dosing** |  |  |  |  |  |
| No vaccine | \$45,459,093,347 | 489,918 | -- | -- | Reference |
| mRNA-1273 | \$48,247,894,967 | 462,594 | \$2,788,801,620 | 27,324 | \$102,065 |
| **Base-Case for the 65+ years (All) Subgroup: Annual dosing** |  |  |  |  |  |
| No vaccine | \$29,081,412,908 | 545,887 | -- | -- | Reference |
| mRNA-1273 | \$29,371,536,897 | 446,364 | \$290,123,989 | 99,522 | \$2,915 |
| **Scenario Analysis: Annual dosing for 6 months-64 years High-Risk, Semi-annual dosing for 65+ All** |  |  |  |  |  |
| No vaccine | \$78,627,073,010 | 1,127,731 | -- | -- | Reference |
| mRNA-1273 | \$82,398,232,261 | 996,069 | \$3,771,159,251 | 131,662 | \$28,643 |

Abbreviations: Δ, difference; ICER, incremental cost-effectiveness ratio; QALY, quality-adjusted life-year
\*Difference in QALYs gained is equivalent to negative one times the difference in total QALYs lost.

**Table 4.** Scenario analysis results (mRNA-1273 compared to no vaccine in the target population)

| Model Parameter | Range/Data Selections | ICER (Δ Cost per QALY Gained)) | % Change from Base-Case |
| --- | --- | --- | --- |
| Base-Case | -- | \$23,265 | Reference |
| Perspective | Healthcare | \$38,698 | 66.3% |
| Premature mortality productivity losses | Excludes both market and non-market productivity losses | \$35,781 | 53.8% |
| Vaccine coverage rates | Increase coverage by 10% for all ages | \$24,376 | 4.8% |
| Vaccine coverage rates | Increase coverage by 5% for all ages | \$23,903 | 2.7% |
| Vaccine coverage rates | Increase coverage by 5% for 18+ only | \$23,818 | 2.4% |
| Vaccine cost | Double the unit cost of mRNA-1273 for ages 6 months – 4 years* | \$23,646 | 1.6% |
| Percentage hospitalized (high-risk) | Increased RR of hospitalization (RR=1.45) for high-risk (18-64 years only) | \$23,561 | 1.3% |
| Inpatient mortality (high-risk) | Increased mortality (RR=1.62) for high-risk (18-64 years only) | \$23,513 | 1.1% |
| Percentage with outpatient care (high-risk) | Assume equal to general population | \$23,474 | 0.9% |
| Caregiver productivity losses | Excluded | \$22,951 | -1.4% |
| Indirect benefit | Calculated based on CDC Right Study | \$21,163 | -9.0% |
| Opportunity costs | Include opportunity costs | \$21,127 | -9.2% |
| VE | VE against symptomatic COVID-19 infection for adults ages 18 years and older from ICATT study | \$17,117 | -26.4% |
| Percentage with no formal care | Based on influenza (2023 - 2024) | \$15,877 | -31.76% |
| Premature mortality productivity losses | Includes both market and non-market productivity losses | \$6,336 | -72.8% |

Abbreviations: Δ, difference; ICER, incremental cost-effectiveness ratio; QALY, quality-adjusted life-year; RR: relative risk; VE: vaccine effectiveness
\*This analysis was also performed for the 6 month – 4 year high-risk subgroup only; doubling the vaccine cost in this subgroup led to mRNA-1273 dominating no vaccine (no change from the base-case).

### Deterministic Sensitivity and Scenario Analyses (mRNA-1273 compared to no vaccine)

The impact of the DSAs and scenario analyses on the number of symptomatic infections, hospitalizations, and deaths prevented are summarized in the Technical Appendix (Section 13). The impact of key DSAs on cost-effectiveness results is presented in Figure 1 (details in Supplemental Table 27), while the scenario analyses are shown below in Table 4.

**Figure 1:**
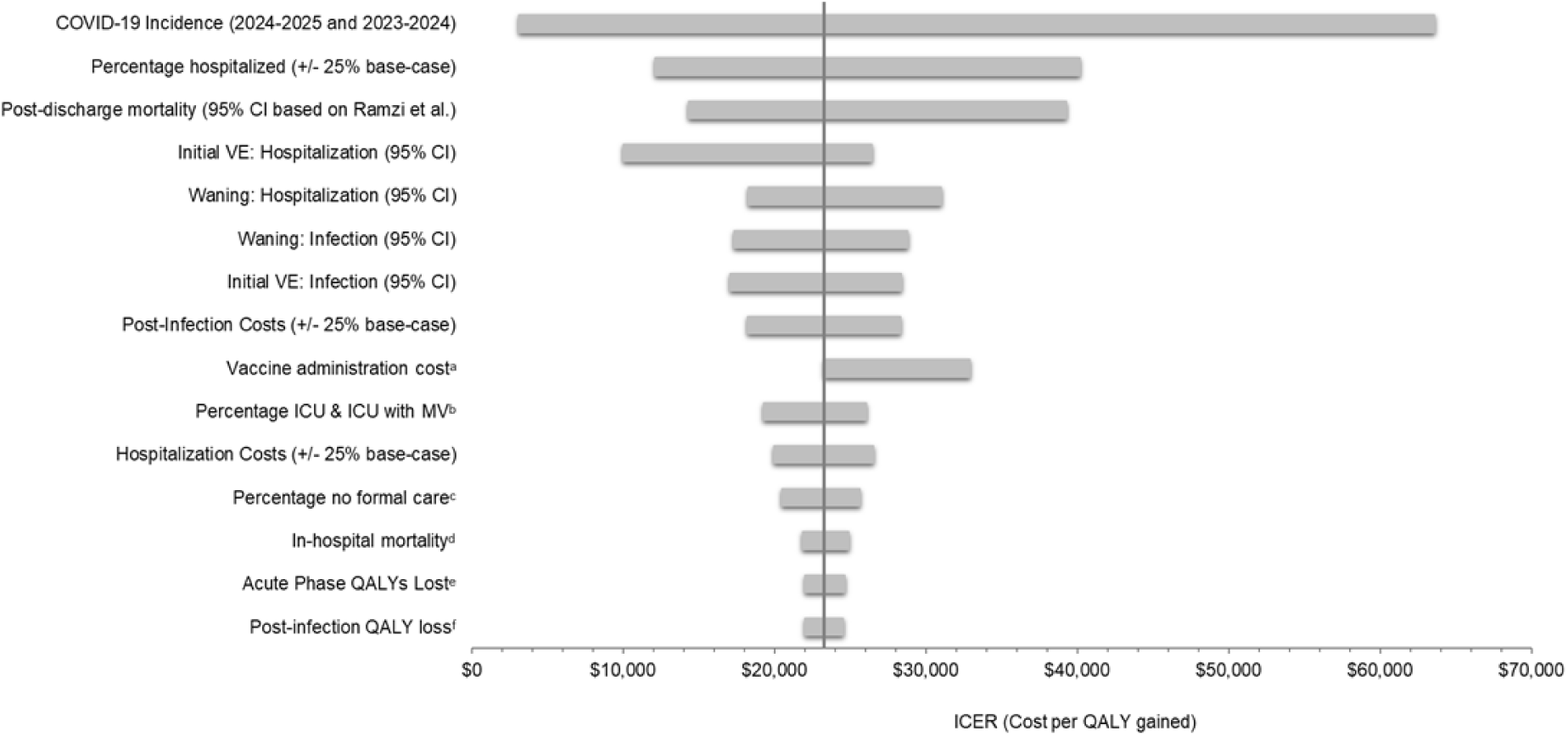
Tornados: Impact of deterministic sensitivity analyses on the base-case cost-effectiveness results (mRNA-1273 relative to no vaccine)

In the DSAs, varying the COVID-19 incidence has the greatest impact on the ICER. Decreasing COVID-19 to estimates based on the 2024-2025 season alone yields an ICER of $63,587 per QALY gained relative to no vaccine (173.3% change from the base-case ICER), while increasing to estimates based on the 2023-2024 season decreases this ICER to $3,148 per QALY gained.

Varying the proportion of patients hospitalized also has a large impact on the ICER; varying to - 25% and +25% of the base-case values yields ICERs of $40,177 (72.3%) and $12,139 (-47.8%) per QALY gained, respectively. Varying post-discharge mortality to the 95% CI upper and lower bounds produces ICERs of $14,352 (38.3% decrease) and $39,272 (68.8% increase) respectively. Finally, varying the mRNA-1273 initial VE against hospitalization based on the 95% CI lower and upper bounds leads to ICERs of $26,437 (13.6% increase) and $9,997 (57.0% decrease), respectively. Other key variables impacting the cost-effectiveness results by >20% include the vaccine administration cost, the mRNA-1273 waning rates against infection and hospitalization, mRNA-1273 initial VE against infection, post-discharge costs, and hospitalization costs.

The scenario analysis performed from the healthcare perspective (excluding both patient and caregiver lost productivity) yielded an ICER of $38,698 per QALY gained, representing a 66.3% increase from the base-case. Including both non-market and market productivity losses associated with premature mortality instead of market productivity losses alone as in the base case reduced the ICER to $6,336 per QALY gained (a 72.8% reduction). Excluding all premature mortality productivity losses leads to a 53.8% increase in the ICER to $35,781 per QALY gained. Increasing the proportion of patients with no formal care for COVID-19 related-illness according to CDC burden of illness influenza data leads to a 31.8% reduction in the ICER. Using data from the ICATT study on the VE against symptomatic infection for adults ≥18 years leads to a 26.4% reduction in the ICER to $17,117 per QALY gained. All other scenario analyses impacted the ICER <10% relative to the base-case.

### Comparison of mRNA-1273 to BNT162b2

Clinical outcomes for the comparison between mRNA-1273 and BNT162b2 are presented in the Technical Appendix (Supplemental Table 25). Considering high-risk adults ages 18-64 and all ≥65 years, mRNA-1273 is expected to avert 539,125 symptomatic infections, 38,476 COVID-19 related hospitalizations and 5,067 deaths compared to BNT162b2. Prevention of these clinical outcomes leads to fewer QALYs lost. As the unit cost of mRNA-1273 is lower than BNT162b2, mRNA-1273 dominates in the base-case analysis (Table 5), all subgroup analyses (Table 5) and all scenario analyses (Supplemental Tables 29 and 30).

**Table 5:**
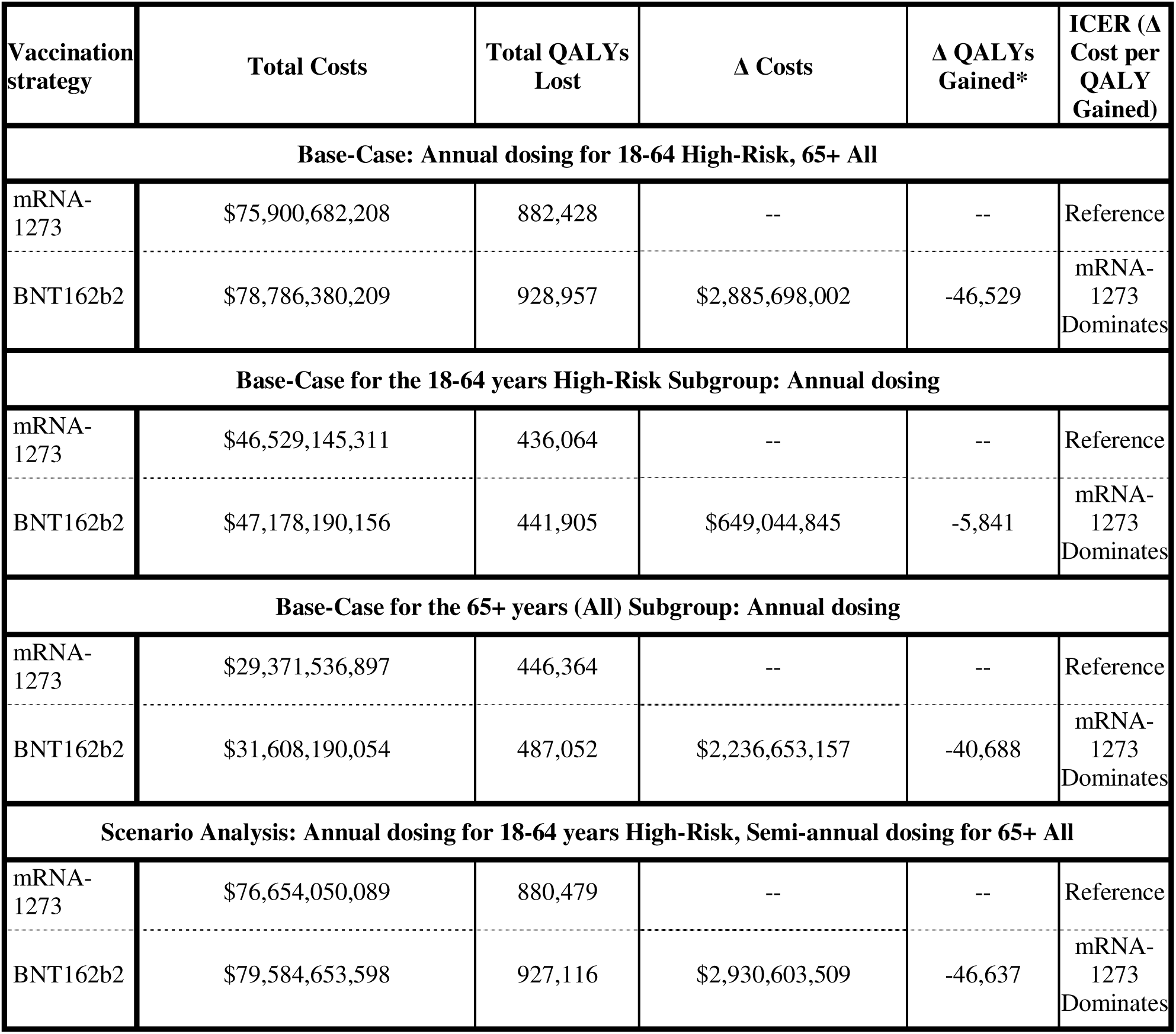
Cost-effectiveness results for mRNA-1273 compared to BNT162b2.

## Discussion

During the 2025/2026 season in the US, we estimate that mRNA-1273 vaccine would prevent, in its licensed target population (adults aged ≥65 years and individuals aged 6 months–64 years with underlying medical conditions), approximately 1.9 million symptomatic COVID-19 infections, 101,739 hospitalizations, and 13,285 deaths compared to no vaccination. Given this clinical impact, annual dosing with mRNA-1273 (base-case analysis) is expected to lead to a gain of 129,712 QALYs with incremental costs of $3.0 billion compared to no vaccine, yielding an ICER (cost per QALY gained) of $23,265 for mRNA-1273 compared to no vaccine. In the subgroups of high-risk children ages 6 months-4 years, mRNA-1273 was shown to dominate no vaccine. In other age sub-groups, ICERs considered cost-effective using a $150,000 per QALY gained willingness-to-pay threshold were estimated ($102,065 for high-risk 5 to 64 year olds; $2,915 for ≥65 years). In the base-case, for every 1 USD of mRNA-1273 vaccine related costs, there is a return of 1.91 to 7.90 dollars in terms of costs from the societal perspective and monetized health benefit gained by preventing SARS-CoV-2 infections and related COVID-19 outcomes.

Compared to BNT162b2, we estimate that mRNA-1273 would have averted 539,125 symptomatic infections, 38,476 hospitalizations, and 5,067 deaths in high-risk adults ages 18-64 years and all adults ≥65 years. Given these additional clinical outcomes averted, combined with a lower vaccine unit cost, mRNA-1273 was shown to dominate BNT162b2.

The results of all sensitivity and scenario analyses suggest that mRNA-1273 still represents a valuable vaccination option—particularly for individuals aged 6 months to 4 years with high-risk medical conditions who remain most vulnerable to severe disease. mRNA-1273 is currently the only licensed vaccine for children ages 6 months – 4 years at high-risk of severe COVID-19 outcomes.^2^ Due to a lack of data capturing dosing patterns in children ages 6 months to 4 years over a 1-year time frame, it is difficult to distinguish the number of children who require a priming 2-dose series versus those requiring a booster dose. In a scenario analysis, the vaccine cost was doubled for those 6 months-4 years only as a proxy for the receipt of two doses of mRNA-1273; when the unit cost of mRNA-1273 was doubled, there was no change from the base-case result of mRNA-1273 dominating no vaccine in this subgroup. Results of this scenario analysis suggest it is highly cost-effective to consider priming with 2 doses in the 6 months-4 years only subgroup.

A recent publication by Prosser et al. (2025) assessed the cost-effectiveness of a 2023-2024 mRNA COVID-19 vaccine with no updated vaccination in adults ages ≥18 years of age.^7^ It is challenging to perform a direct comparison between Prosser et al. (2025) and the present study due to differences in model design and presentation of results. For example, Prosser et al. report ICERs by age group, whereas the emphasis of the present study is to report ICERs for the fully licensed population for mRNA-1273. There are also key differences between studies in incidence estimates and VE. The present study estimates fewer symptomatic infections for the same number of hospitalizations compared to Prosser et al., yielding a more conservative estimate of the number of symptomatic infections. Further, the present study uses VE estimates specific to mRNA-1273, where available, which are higher than the VE estimates reported by Prosser et al. The limitations discussed in the previous manuscript by Fust et al. also apply to this analysis. For example, the differences between mRNA-1273 and BNT-162b2 are based on indirect meta-analyses and these results should be confirmed in a direct comparison of the vaccines. However, the unit cost of mRNA-1273 is lower than BNT-162b2 and therefore all of the comparative economic analyses favored mRNA-1273. In addition, in the modeled population <18 years of age, there are limited data related to the long-term outcomes of COVID-19 infection. Further, the risk factors for COVID-19 related hospitalization in the pediatric population are heterogenous across age subgroups; for example, conditions such as prematurity may be a risk factor for COVID-19 related hospitalization for those <2 years of age, while conditions such as neurologic disorders and asthma/reactive airway disease may play a larger role in COVID-19 related hospitalizations for children ages 2-17 years.^30^ Data were also limited for the pediatric population related to high-risk hospitalization and outpatient care cost estimates, as well as in-hospital mortality stratified by in-hospital setting of care. Finally, it is difficult to distinguish the proportion of patients ages 6 months – 4 years receiving a booster dose versus the primary series; however, results of a scenario analysis assuming double the cost of vaccination for all ages 6 months – 4 years was performed and demonstrated that mRNA-1273 was dominant compared to no vaccine, indicating that even if 100% of those in this age group required a priming series, mRNA-1273 would still represent a cost-effective option.

## Conclusions

Results of this study suggests that the COVID-19 mRNA-1273 vaccine could substantially reduce the clinical and economic burden of COVID-19 among all patients who may receive a 2025-2026 formulation in the US. These benefits were observed both in comparison to no vaccination and the BNT162b2 vaccine. mRNA-1273 is currently the only licensed vaccine for children ages 6 months – 4 years at high-risk of severe COVID-19 outcomes , and continues to be a highly cost-effective intervention in the rest of the eligible population. As a result of both its indication and demonstrated clinical and economic benefits, mRNA-1273 continues to be an important part of the overall COVID-19 vaccination program.

### Transparency

- **Declaration of Funding:** This study was supported by Moderna, Inc.
- **Declaration of competing interests:** MK is a shareholder in Quadrant Health Economics Inc, which was contracted by Moderna, Inc. to conduct this study. KF, MW and SC are consultants to Quadrant Health Economics Inc. KJ, NV, and EB are employed by Moderna, Inc. and may hold stock/stock options in the company.

## Data Availability

All data produced in the present work are contained in the manuscript.

