## Supplement for "Modeling the Cost-Effectiveness of the COVID-19 mRNA-1273 vaccine in the United States"

The Potential Clinical Impact of the COVID-19 mRNA-1273 Vaccine

- Technical Appendix –

### 1 Estimation of Incidence of Infection (No Vaccination)

As the hospitalization data from the CDC reflects a partially vaccinated population, information on vaccination coverage, VE, and the probability of hospitalization must be used as well as the target hospitalization rates. Two calibrations were completed, the first using data from September 2023 to August 2024 and the second for September 2024 to August 2025. The process is described below.

**Step 1**: Develop the age-specific targets for the monthly rate of hospitalizations by age group for a one-year period. As hospitalization rates for one full year, i.e. 12 months, are needed, for the base case, the hospitalization rates for September 2023 to August 2024 and September 2024 to August 2025 from COVID-NET^[1]^ were used.^1^ The rates per 100,000 are displayed in Table 1 and Table 2 below.

Table 1. Target hospitalization rates per 100,000 for the calculation of infection incidence in the model (September 2023 – August 2024)^1^

| **Age Group (Years)** | **Sep-23** | **Oct-23** | **Nov-23** | **Dec-23** | **Jan-24** | **Feb-24** | **Mar-24** | **Apr-24** | **May-24** | **Jun-24** | **Jul-24** | **Aug-24** |
| --- | --- | --- | --- | --- | --- | --- | --- | --- | --- | --- | --- | --- |
| 0-4 | 9.3 | 9.3 | 13.2 | 18.7 | 18.2 | 11.6 | 6.4 | 3.6 | 3.5 | 4.4 | 9 | 12.1 |
| 5-17 | 1.6 | 0.9 | 1.4 | 2.2 | 2.6 | 2.4 | 1 | 0.5 | 0.6 | 0.3 | 1.2 | 1.6 |
| 18-49 | 5.5 | 5.2 | 6.1 | 8.3 | 9 | 5.7 | 2.9 | 1.6 | 1.6 | 2.6 | 5.3 | 6.7 |
| 50-64 | 15.9 | 15.2 | 18.8 | 24 | 26.4 | 16.2 | 8.8 | 5 | 4.4 | 6.4 | 13 | 17.2 |
| ≥ 65 | 80.9 | 80.7 | 91.1 | 123.2 | 113.3 | 68.8 | 43 | 25.7 | 24.4 | 39.3 | 66.5 | 82.5 |

Table 2. Target hospitalization rates per 100,000 for the calculation of infection incidence in the model (September 2024 – August 2025)^1^

| **Age Group (Years)** | **Sep-24** | **Oct-24** | **Nov-24** | **Dec-24** | **Jan-25** | **Feb-25** | **Mar-25** | **Apr-25** | **May-25** | **Jun-25** | **Jul-25** | **Aug-25** |
| --- | --- | --- | --- | --- | --- | --- | --- | --- | --- | --- | --- | --- |
| 0-4 | 8.4 | 5.6 | 4.1 | 6.1 | 9 | 6.2 | 5.1 | 3.4 | 2.9 | 2 | 4.3 | 7.8 |
| 5-17 | 1.6 | 0.9 | 0.6 | 0.9 | 1.2 | 0.8 | 0.7 | 0.5 | 0.3 | 0.3 | 0.5 | 1.1 |
| 18-49 | 4.3 | 2.6 | 1.9 | 3.1 | 3.9 | 2.9 | 2.4 | 1.6 | 1.2 | 1.1 | 1.9 | 3.8 |
| 50-64 | 13.2 | 8.7 | 6.2 | 9.1 | 12.5 | 9.1 | 7 | 4.3 | 3 | 2.5 | 4.1 | 7 |
| ≥ 65 | 66.3 | 46.8 | 30.8 | 54.4 | 59.9 | 40.9 | 35.5 | 24.4 | 17.6 | 14.9 | 18.4 | 33.2 |

**Step 2**: Enter the age-specific probability of hospitalization and the related proportion of symptomatic cases that are seeking care. The same probabilities are used for the cost-effectiveness model of the symptomatic infection in the no vaccination arm and the incidence calculation.

**Step 3**: Enter the assumed vaccination coverage.

The monthly vaccine coverage rates for September 2023 to August 2024 (Table 3) and September 2024 to August 2025 were estimated from the vaccine coverage from the CDC VaxView database.^2,3^ Data on coverage of a second dose of the vaccine were also available for individuals aged 65 years and older, and began April 1, 2024 and end July 27, 2024. Figure 1 provides detailed vaccine coverage over time for the second doses.

A small proportion of people aged 65 years and older in the US were vaccinated with a second dose between April and July 2024 (See Figure 1). The model only accommodates one dose for the calculations of infection incidence, so the single dose coverage rates were increased to account for second doses in spring 2024 and spring 2025. Overall, excluding these vaccinations or including them made a small difference to the final infection incidence rates.

Figure 1. Second dose, COVID-19 vaccine coverage during season 2023-2024

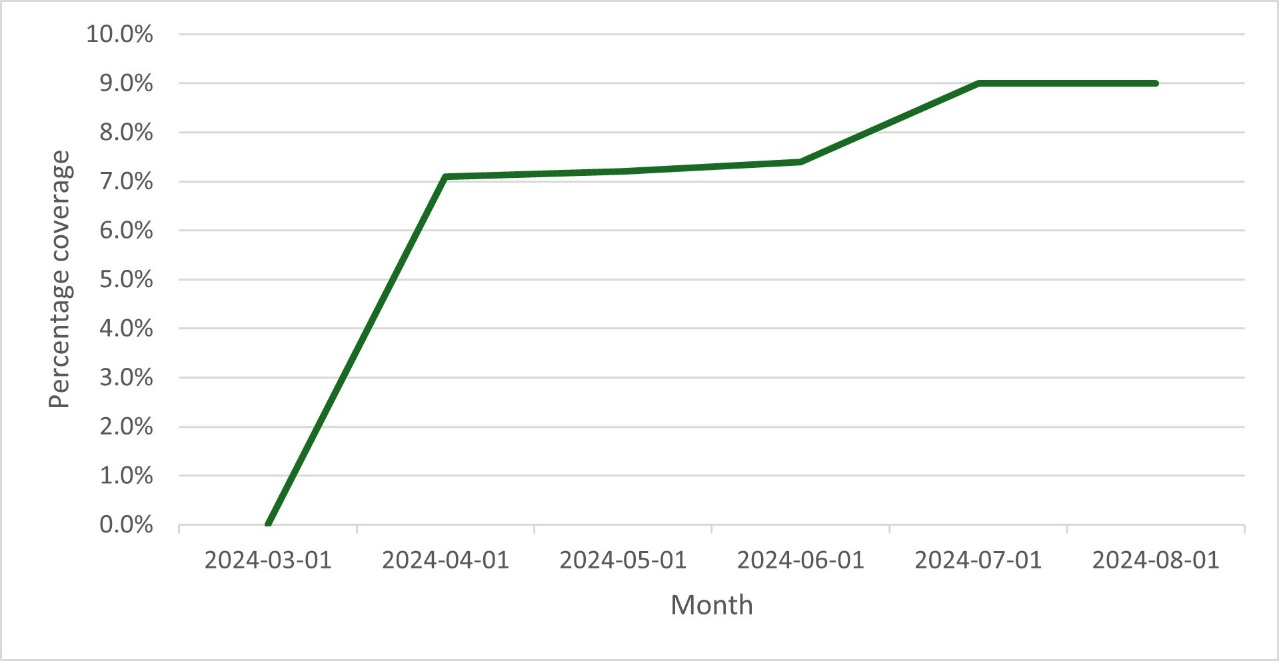

Table 3. Monthly coverage rates (September 2023 – August 2024) used in the calculation of infection incidence in the static model

| **Date** | **0-4 years** | **5-17 years** | **18-49 years** | **50-64 years** | **65+ years** |
| --- | --- | --- | --- | --- | --- |
| 30-Sep-2023 | 0.2% | 1.1% | 1.4% | 3.9% | 6.1% |
| 31-Oct-2023 | 1.6% | 6.9% | 5.6% | 12.3% | 22.5% |
| 30-Nov-2023 | 3.8% | 11.3% | 9.7% | 18.0% | 28.9% |
| 31-Dec-2023 | 4.3% | 12.7% | 11.3% | 20.7% | 32.2% |
| 31-Jan-2024 | 5.4% | 14.6% | 12.9% | 22.7% | 35.1% |
| 29-Feb-2024 | 5.7% | 15.5% | 13.5% | 23.4% | 36.1% |
| 31-Mar-2024 | 6.0% | 16.2% | 13.9% | 23.7% | 37.7% |
| 30-Apr-2024 | 6.5% | 16.6% | 14.2% | 24.2% | 45.4% |
| 31-May-2024 | 6.7% | 17.2% | 14.3% | 24.3% | 45.8% |
| 30-Jun-2024 | 6.7% | 17.3% | 14.3% | 24.3% | 46.0% |
| 31-Jul-2024 | 6.7% | 17.3% | 14.3% | 24.3% | 47.6% |
| 31-Aug-2024 | 6.7% | 17.3% | 14.3% | 24.3% | 47.6% |

Table 4. Monthly coverage rates (September 2024 – August 2025) used in the calculation of infection incidence in the static model

| **Date** | **0-4 years** | **5-17 years** | **18-49 years** | **50-64 years** | **65+ years** |
| --- | --- | --- | --- | --- | --- |
| 30-Sep-2024 | 1.0% | 3.6% | 3.3% | 7.5% | 15.8% |
| 31-Oct-2024 | 2.8% | 8.5% | 8.4% | 15.7% | 31.2% |
| 30-Nov-2024 | 4.1% | 10.8% | 10.9% | 20.3% | 38.0% |
| 31-Dec-2024 | 4.7% | 12.8% | 12.5% | 22.8% | 42.2% |
| 31-Jan-2025 | 5.0% | 13.8% | 13.6% | 24.0% | 44.0% |
| 29-Feb-2025 | 5.2% | 14.6% | 14.1% | 24.8% | 44.4% |
| 31-Mar-2025 | 5.4% | 14.9% | 14.1% | 24.8% | 44.4% |
| 30-Apr-2025 | 5.6% | 15.4% | 14.1% | 24.8% | 51.5% |
| 31-May-2025 | 5.6% | 15.4% | 14.1% | 24.8% | 51.6% |
| 30-Jun-2025 | 5.6% | 15.4% | 14.1% | 24.8% | 51.8% |
| 31-Jul-2025 | 5.6% | 15.4% | 14.1% | 24.8% | 53.4% |
| 31-Aug-2025 | 5.6% | 15.4% | 14.1% | 24.8% | 53.4% |

**Step 4**: Enter the assumed initial vaccine effectiveness (VE) against infection and hospitalization and the monthly linear waning rate over time. **See Section 2.2**.

In the model calculation sheets, an initial set of monthly incidence rates of symptomatic infection without seasonal vaccination is used to calculate the number of hospitalizations by month and age group. These estimated hospitalization counts are compared to the target number of hospitalizations by month and age group and the ratio of target to model estimated hospitalizations is calculated for each month and age group. These ratios are multiplied by the initial set of monthly incidence rates to determine the calibrated monthly incidence rates by age group that are required to estimate the target number of hospitalizations, given the other model inputs are held constant. These final incidence rates are then output and can be copied directly into the static CEA model. The base case incidence values are displayed in Table 5 (2023-24) and Table 6 (2024-25). The average values of 2023-24 and 2024-25, as displayed in Table 7, were used for the base case.

Table 5. 2023-24 incidence of symptomatic infection (no vaccination arm) for the static model (% infected)

| **Age group (years)** | **Sep-23** | **Oct-23** | **Nov-23** | **Dec-23** | **Jan-24** | **Feb-24** | **Mar-24** | **Apr-24** | **May-24** | **Jun-24** | **Jul-24** | **Aug-24** |
| --- | --- | --- | --- | --- | --- | --- | --- | --- | --- | --- | --- | --- |
| 0-4 | 5.2% | 5.2% | 7.5% | 10.7% | 10.5% | 6.7% | 3.7% | 2.1% | 2.0% | 2.5% | 5.2% | 6.9% |
| 5-17 | 0.9% | 0.5% | 0.8% | 1.3% | 1.6% | 1.4% | 0.6% | 0.3% | 0.4% | 0.2% | 0.7% | 0.9% |
| 18-49 | 2.0% | 1.9% | 2.3% | 3.1% | 3.4% | 2.2% | 1.1% | 0.6% | 0.6% | 1.0% | 2.0% | 2.5% |
| 50-64 | 1.4% | 1.5% | 1.9% | 2.4% | 2.7% | 1.6% | 0.9% | 0.5% | 0.4% | 0.6% | 1.3% | 1.7% |
| 65+ | 1.0% | 1.1% | 1.3% | 1.8% | 1.7% | 1.0% | 0.6% | 0.4% | 0.4% | 0.6% | 1.0% | 1.3% |

Table 6. 2024-25 incidence of symptomatic infection (no vaccination arm) for the static model (% infected)

| **Age group (years)** | **Sep-24** | **Oct-24** | **Nov-24** | **Dec-24** | **Jan-25** | **Feb-25** | **Mar-25** | **Apr-25** | **May-25** | **Jun-25** | **Jul-25** | **Aug-25** |
| --- | --- | --- | --- | --- | --- | --- | --- | --- | --- | --- | --- | --- |
| 0-4 | 4.7% | 3.2% | 2.4% | 3.5% | 5.2% | 3.6% | 3.0% | 2.0% | 1.7% | 1.2% | 2.5% | 4.5% |
| 5-17 | 0.9% | 0.5% | 0.3% | 0.5% | 0.7% | 0.5% | 0.4% | 0.3% | 0.2% | 0.2% | 0.3% | 0.6% |
| 18-49 | 1.5% | 1.0% | 0.7% | 1.2% | 1.5% | 1.1% | 0.9% | 0.6% | 0.4% | 0.4% | 0.7% | 1.4% |
| 50-64 | 1.2% | 0.8% | 0.6% | 0.9% | 1.3% | 0.9% | 0.7% | 0.4% | 0.3% | 0.2% | 0.4% | 0.7% |
| 65+ | 0.9% | 0.7% | 0.5% | 0.8% | 0.9% | 0.6% | 0.5% | 0.4% | 0.3% | 0.2% | 0.3% | 0.5% |

Table 7. Base case incidence of symptomatic infection (no vaccination arm) for the static model (% infected) (Average of 2023-2024 and 2024-2025 seasons)

| **Age group (years)** | **Sep** | **Oct** | **Nov** | **Dec** | **Jan** | **Feb** | **Mar** | **Apr** | **May** | **Jun** | **Jul** | **Aug** |
| --- | --- | --- | --- | --- | --- | --- | --- | --- | --- | --- | --- | --- |
| 0-4 | 4.9% | 4.2% | 4.9% | 7.1% | 7.8% | 5.1% | 3.3% | 2.0% | 1.8% | 1.8% | 3.8% | 5.7% |
| 5-17 | 0.9% | 0.5% | 0.6% | 0.9% | 1.1% | 1.0% | 0.5% | 0.3% | 0.3% | 0.2% | 0.5% | 0.8% |
| 18-49 | 1.8% | 1.4% | 1.5% | 2.2% | 2.4% | 1.6% | 1.0% | 0.6% | 0.5% | 0.7% | 1.3% | 2.0% |
| 50-64 | 1.3% | 1.1% | 1.2% | 1.7% | 2.0% | 1.3% | 0.8% | 0.5% | 0.4% | 0.4% | 0.8% | 1.2% |
| 65+ | 0.9% | 0.9% | 0.9% | 1.3% | 1.3% | 0.8% | 0.6% | 0.4% | 0.3% | 0.4% | 0.6% | 0.9% |

### 2 Vaccine Effectiveness

#### 2.1 Calculation of Vaccine Effectiveness with the Model

Vaccine effectiveness declines linearly on a monthly basis within the model. A portion of the cohort can receive a new vaccine in each month of the time horizon. In order to incorporate waning of effectiveness, the effectiveness calculation is a function of the fraction of the age group vaccinated each month and vaccine effectiveness. Vaccine effectiveness is the sum of the following:

$Effectiveness={Cov}_{Cur}\times{Eff}_{Cur}+{Cov}_{Cur-1}\times{Eff}_{Cur-1}+ {Cov}_{Cur-2}\times{Eff}_{Cur-2}+\ldots+ {Cov}_{Cur-9}\times{Eff}_{Cur-9}$

Where:

Cov_cur_ = fraction of age group vaccinated in the current month

Cov_cur1_ = fraction of age group vaccinated in the month before the current month

Cov_cur2_ = fraction of the age group vaccinated two months before the current month

Eff_cur_ = effectiveness in the current month

Eff_cur1_ = effectiveness one month after initial vaccination

Eff_cur2_ = effectiveness two months after initial vaccination

#### 2.2 Inputs for the Estimation of Incidence

The VE inputs used for the derivation of incidence are shown below. Please see the Appendix in the article by Fust et al for more information.^4^ In the original article, the pediatric population was limited to those 12 to 17 years but the CDC publications used for 2023-24^5^ and 2024-25^6^ also contained data for all age pediatric age groups. Therefore these studies were used to estimate the VE for all COVID-19 vaccines for the 0-4 year and 5-17 year age groups.

Table 8. Market mix VE used in the calculation of infection incidence in the model (2023/24 season)

| **Age Group** | **Infection** | | **Hospitalization** | |
| --- | --- | --- | --- | --- |
|  | **Initial VE** | **Waning** | **Initial VE** | **Waning** |
| 0 to 4 years | 0.66 | 0.0475 | 0.66 | 0.0246 |
| 5 to 17 years | 0.71 | 0.0475 | 0.71 | 0.0246 |
| 18-49 years | 0.402 | 0.0475 | 0.627 | 0.0246 |
| 50-64 years | 0.402 | 0.0475 | 0.627 | 0.0246 |
| ≥65 years | 0.402 | 0.0475 | 0.627 | 0.0246 |

VE: Vaccine effectiveness

Table 9. Market mix VE used in the calculation of infection incidence in the model (2024/25 season)

| **Age Group** | **Infection** | | **Hospitalization** | |
| --- | --- | --- | --- | --- |
|  | **Initial VE** | **Waning** | **Initial VE** | **Waning** |
| 0 to 4 years | 0.844 | 0.0475 | 0.844 | 0.0246 |
| 5 to 17 years | 0.624 | 0.0475 | 0.624 | 0.0246 |
| 18-49 years | 0.457 | 0.0475 | 0.544 | 0.0246 |
| 50-64 years | 0.457 | 0.0475 | 0.544 | 0.0246 |
| ≥65 years | 0.457 | 0.0475 | 0.544 | 0.0246 |

VE: Vaccine effectiveness

#### 2.3 Calculation of Incremental VE against hospitalization

The model assumes the VE against infection to be lower than the VE against hospitalization and thus applies an incremental VE against hospitalization for infections occurring in vaccinated in the vaccination arm. The hospitalization VE values are adjusted in the model to reflect the incremental protection against hospitalization above the protection against infection. In other words, VE values were adjusted to account for cases of hospitalization that are prevented due to the decrease in infections with vaccinations. This was to ensure that protection against hospitalizations is not double counted. The equations used for this are presented below.

For each age group, we define the following vaccine effectiveness variables and relationship between the variables. The superscripts and subscripts for age group are removed for clarity.

**Definitions**

${VE}_{1}$ = Vaccine effectiveness against infection

${VE}_{2}$ = ‘Total’ Vaccine effectiveness against hospitalization

${VE}_{2}^{*}$= ‘Additional’ Vaccine effectiveness against hospitalization

We assume ${VE}_{2}^{*}=0$ if there is no additional benefit against hospitalization

**Define**

$$\left[ 1-{VE}_{2} \right]= \left[ 1-{VE}_{1} \right]\times\left[ 1-{VE}_{2}^{*} \right]$$

Isolate and solve for ${VE}_{2}^{*}$

$$\left[ 1-{VE}_{2}^{*} \right]=\frac{\left[ 1-{VE}_{2} \right]}{\left[ 1-{VE}_{1} \right]}$$

$${VE}_{2}^{*}= 1- \frac{\left[ 1-{VE}_{2} \right]}{\left[ 1-{VE}_{1} \right]}$$

### 3 Cohort Size

Table 10. Cohort eligible for vaccination

| **Age Group (Years)** | **Population Size** | **Percent Eligible*** | **Eligible Population Size** | **Source** |
| --- | --- | --- | --- | --- |
| 6 mos – 4 | 15,078,009 | 45.7% | 6,890,650 | United Nations (2024)^7^; Forrest (2025)^8^ |
| 5 - 17 | 55,468,504 | 45.7% | 25,349,106 | United Nations (2024)^7^; Forrest (2025)^8^ |
| 18-49 | 145,627,454 | 65.7% | 95,738,794 | United Nations (2024)^7^; Panagiotakopoulos (2025)^9^ |
| 50-64 | 63,863,350 | 81.0% | 51,729,314 |  |
| ≥65 | 59,874,349 | 100% | 59,874,349 | United Nations (2024)^7^ |

*Reflects the proportion considered high-risk for ages 6 months-64 years; for ages 65+, all adults are eligible for vaccination.

*High-risk conditions for severe COVID-19 among those 3-17 years studied by Forrest et al. (2025) includes 97 chronic conditions based on clinical reported data from PEDSnet.

^†^High-risk conditions for those 18+ years reflect those included in Panagiotakopoulous et al., and include asthma, cancer (hematologic malignancies), cerebrovascular disease, chronic kidney disease (dialysis), chronic lung diseases (bronchiectasis, COPD, interstitial lung disease, pulmonary embolism, pulmonary hypertension), chronic liver diseases (cirrhosis, non-alcoholic fatty liver disease, alcoholic liver disease, autoimmune hepatitis), cystic fibrosis, diabetes mellitus (types 1 and 2), disabilities, heart conditions, HIV, mental health conditions (depression, schizophrenia spectrum disorders), neurologic conditions (dementia; Parkinson’s disease), obesity, physical inactivity, pregnancy and recent pregnancy, primary immunodeficiencies, smoking (current and former), solid organ or blood stem cell transplantation, tuberculosis, use of corticosteroids or other immunosuppressive medications

### 4 Vaccine Coverage

A figure showing the vaccine uptake by month used for the analytic time horizon, based on data from September 2024 to February 2025 from COVIDVaxView,^2,3^ is displayed below. The uptake of the second dose for the scenario where semi-annual vaccination of those 65 years and older is tested is shown in Section 1 (Figure 2).

Based on observed increases of CDC reported COVID-19 vaccination coverage rates in persons 65 years and older between season 2023/2024 and 2024/2025, additional scenarios for 1-dose vaccination strategies were studied assuming an increase in vaccination coverage rates by 5% and 10% in the overall population (i.e., 6 months to 64 years high-risk and 65 years and older); an additional analysis assuming a coverage increase of 5% in those ≥18 years was also performed. The corresponding vaccination uptake is shown in Table 11.

Figure 2. First seasonal dose vaccine coverage by age used for the analytic time horizon.

Percent Coverage

|  | 0-4 years | 5-17 years | 18-49 years | 50-64 years | 65+ years |
| --- | --- | --- | --- | --- | --- |
| Sep | 0.984806% | 3.60385% | 3.3% | 7.5% | 15.8% |
| Oct | 2.83133% | 8.50794% | 8.4% | 15.7% | 31.2% |
| Nov | 4.1121% | 10.8047% | 10.9% | 20.3% | 38% |
| Dec | 4.6886% | 12.7966% | 12.5% | 22.8% | 42.2% |
| Jan | 4.99199% | 13.792% | 13.6% | 24% | 44% |
| Feb | 5.2335% | 14.6047% | 14.1% | 24.8% | 44.4% |
| Mar | 5.35629% | 14.9006% | 14.1% | 24.8% | 44.4% |
| Apr | 5.5979% | 15.3575% | 14.1% | 24.8% | 51.5% |
| May | 5.5979% | 15.3575% | 14.1% | 24.8% | 51.6% |
| Jun | 5.5979% | 15.3575% | 14.1% | 24.8% | 51.8% |
| July | 5.5979% | 15.3575% | 14.1% | 24.8% | 53.4% |
| Aug | 5.5979% | 15.3575% | 14.1% | 24.8% | 53.4% |

Table 11. Vaccine coverage rates for additional vaccine coverage rate scenarios

|  | **5% increase overall population (6 mos to 64 years high-risk and 65 plus) (1-dose)** | | | | | | **10% increase overall population (6 mos to 64 years high-risk and 65 plus) (1-dose)** | | | | |
| --- | --- | --- | --- | --- | --- | --- | --- | --- | --- | --- | --- |
| **Age group** | **6 mos to 4 years** | **5-17 years** | **18-49 years*** | **50-64 years*** | **65-100 years*** | **6 mos to 4 years** | | **5-17 years** | **18-49 years*** | **50-64 years*** | **65-100 years*** |
| Annual | 5.98% | 8.60% | 8.30% | 12.50% | 20.80% | 10.98% | | 13.60% | 13.30% | 17.50% | 25.80% |
| Sept | 7.83% | 13.51% | 13.40% | 20.70% | 36.20% | 12.83% | | 18.51% | 18.40% | 25.70% | 41.20% |
| Oct | 9.11% | 15.80% | 15.90% | 25.30% | 43.00% | 14.11% | | 20.80% | 20.90% | 30.30% | 48.00% |
| Nov | 9.69% | 17.80% | 17.50% | 27.80% | 47.20% | 14.69% | | 22.80% | 22.50% | 32.80% | 52.20% |
| Dec | 9.99% | 18.79% | 18.60% | 29.00% | 49.00% | 14.99% | | 23.79% | 23.60% | 34.00% | 54.00% |
| Jan | 10.23% | 19.60% | 19.10% | 29.80% | 49.40% | 15.23% | | 24.60% | 24.10% | 34.80% | 54.40% |
| Feb | 10.36% | 19.90% | 19.10% | 29.80% | 49.40% | 15.36% | | 24.90% | 24.10% | 34.80% | 54.40% |
| March | 10.60% | 20.36% | 19.10% | 29.80% | 56.50% | 15.60% | | 25.36% | 24.10% | 34.80% | 61.50% |
| April | 10.60% | 20.36% | 19.10% | 29.80% | 56.60% | 15.60% | | 25.36% | 24.10% | 34.80% | 61.60% |
| May | 10.60% | 20.36% | 19.10% | 29.80% | 56.80% | 15.60% | | 25.36% | 24.10% | 34.80% | 61.80% |
| June | 10.60% | 20.36% | 19.10% | 29.80% | 58.40% | 15.60% | | 25.36% | 24.10% | 34.80% | 63.40% |
| July | 10.60% | 20.36% | 19.10% | 29.80% | 58.40% | 15.60% | | 25.36% | 24.10% | 34.80% | 63.40% |
| August | 5.98% | 8.60% | 8.30% | 12.50% | 20.80% | 10.98% | | 13.60% | 13.30% | 17.50% | 25.80% |

*Estimates also used in an alternative analysis assuming an increase in coverage of 5% for those aged ≥18 years

### 5 Model Probabilities

Most of the model inputs were described in a previous manuscript^4^ and only the values or differences from that manuscript are described below. The model inputs are summarized in Table 12.

Following a symptomatic infection, patients enter the model decision tree and are divided by their highest level of medical care received: 1) No formal health care (not hospitalized nor was outpatient care received); 2) Outpatient care (not hospitalized but did receive outpatient physician visits or emergency department visits without hospital admission); and 3) Inpatient care. The age-specific probabilities of hospitalization given symptomatic infection and the proportion seeking care were estimated based on database analyses and adjusted to represent the high-risk population for ages 6 months to 64 years.

In the decision tree, hospitalized patients were further stratified by location of care, including general ward (no intensive care unit [ICU] or ventilation required), ICU (excluding extracorpeal membrane oxygenation or invasive ventilation), or ICU with mechanical ventilation based on data from the COVID-Net surveillance system.^10^ In absence of data, the proportions for the high-risk populations were assumed to be equivalent to the general population for those ≥18 years. For the high-risk population ages 6 months to 17 years, the in-hospital location of care estimates from the COVID-Net surveillance system^10^ were adjusted using weighted average adjusted relative risks (aRR) calculated based on data from Free et al. (2025),^11^ who report age-specific aRRs by underlying medical condition for severe COVID-19 outcomes including ICU admission and the use of invasive mechanical ventilation. Data presented by Dr. Havers at the April 2025 ACIP meeting^12^ were used to identify the top 4 underlying conditions (chronic lung disease, asthma, neurologic disorders, and obesity) in children ages 2 to 17 years. A weighted average aRR from Free et al. reflecting these 4 conditions was calculated using the relative weights of these conditions based on the ACIP presentation, resulting in aRR estimates for severe COVID-19 outcomes of 1.32 for those ages 2-4 years and 1.30 for those ages 5-17 years (reflecting a weighted average of the 5-11 and 12-17 year-old subgroups). Although data from Free et al. are available for the 6 months to 23 year-old age group, the aRR estimate for those ages 2-4 years was applied to this population, as the conditions for the 2-4 year old age group more accurately reflect the top 4 risk factors identified in the ACIP data for severe COVID-19 for those <18.

Hospitalized patients may die, be discharged and then readmitted, or simply discharged. Estimates of in-hospital mortality for the general population aged ≥5 years^13^, stratified by age and in-hospital location of care, were based on COVID-NET data from March 2022 – October 2023 as analyzed by the University of Michigan COVID-19 Vaccination Modeling Team.^13^ Based on Joshi et al. (2025) ^14^, it was conservatively assumed that the relative risk for in-hospital mortality for high-risk patients aged ≥18 years relative to non-high-risk patients was 1.23, considering an estimate derived for patients with diabetes. In-hospital mortality estimates for those ages 5-17 years were not adjusted by risk status to avoid double-counting (given that the in-hospital location of care estimates are already adjusted for underlying conditions).

The University of Michigan COVID-19 Vaccination Modeling Team^13^ does not provide data for those <5 years; accordingly, data from both the University of Michigan COVID-19 Vaccination Modeling Team^13^ and Andersen et al.^15^ were used to estimate in-hospital mortality rates, by location of care, for the 6 months – 4 years population. Andersen et al.^15^ provide an overall in-hospital mortality estimate (i.e., data are not stratified by in-hospital location of care).  Mortality estimates, by hospital ward, for ages 5-11 years were available from the University of Michigan COVID-19 Vaccination Modeling Team.^13^ These estimates were scaled down to match the overall mortality observed in Andersen et al.^15^ while maintaining the relative values between hospital wards from the University of Michigan COVID-19 Vaccination Modeling Team.^13^

The COVID-NET surveillance system tracks only in-hospital deaths and not post-discharge mortality; similarly, Andersen et al. reports in-hospital death only. As described in Fust et al.^4^, the 30-day readmission and post-discharge mortality probabilities for those ≥18 years were estimated from a meta-analysis for the general population; the readmission estimate employed in the model for those aged ≥18 years (9.35%) represents a weighted average of the high-risk population ages 18-64 and the general population ages ≥65 years.^16^ Estimates of readmission for those <18 years (8.64%) were obtained from Healthcare Cost and Utilization Project (HCUP)^17^ data using diagnosis code INF012 (Coronavirus disease 2-19 [COVID-19]) and reflects the 30-day readmission rate for any cause. Readmission rates for those <18 were not adjusted for risk status, as the in-hospital location of care estimates have already been adjusted for those at high-risk. The resulting readmission rate used in the model (9.20%) reflects a weighted average of the estimates for those < and ≥18 years of age. In the absence of post-discharge mortality specific to those <18 years of age, the 7.87% (95% CI: 2.78%, 12.96%)^16^ used for those ≥18 years was utilized.

The probability of long COVID did not change from the previous manuscript as it affected those ≥18 years only. All patients with COVID-19 infection are subject to risk of infection-induced myocarditis, which is applied as a toll. These probabilities also did not change from the previous manuscript.

Table 12. Key decision tree probabilities, base case and range for deterministic sensitivity analyses

| **Parameter** | | **Base (Range)** | | | | **Source** |
| --- | --- | --- | --- | --- | --- | --- |
| **Distribution of Care** | | | | | | |
| **Age Group** | | **% Hospitalized** | **% No Formal Care** | **% Outpatient Care** | |  |
| 6 mos – 4 years (high-risk) | | 0.37% | 94.3%  (92.5%, 94.3%)^b^ | 5.32% | | Kopel et al. (2024) ^18^; Optum’s de-identified Clinformatics® Data Mart Database ^19^; Veradigm (EHR/ claims) database analyses |
| 5-17 years (high-risk) | | 0.12% | 94.3%  (92.5%, 94.3%)^b^ | 5.56% | | Kopel et al. (2024) ^18^; Optum’s de-identified Clinformatics® Data Mart Database ^19^; Veradigm (EHR/ claims) database analyses |
| 18-49 years (high-risk) | | 0.33%  (0.25%, 0.41%) ^a^ | 90.0%  (88.9%, 90.0%)^b^ | 9.67% | | Kopel et al. (2024) ^18^; Veradigm (EHR/claims) database analyses^20^ |
| 50-64 years (high-risk) | | 1.22%  (0.92%, 1.53%) ^a^ | 73.9%  (73.0%, 73.9%)^b^ | 24.88% | |  |
| 65+ years (all) | | 8.29%  (6.22%, 10.36%) ^a^ | 38.5%  (28.9%, 48.1%)^a^ | 53.2% | | Kopel et al. (2024)^18^ |
| **Location of Care for Hospitalized Patients*** | | | | | | |
| **Age Group** | | **No ICU or MV** | **ICU Only** | **ICU with MV** | |  |
| 6 months – 4 years (high-risk) | | 72.54% | 19.45% | 8.01% | | COVID-Net^10^; Free et al. (2025)^11^ |
| 5-17 years (high-risk) | | 72.74% | 19.30% | 7.95% | | COVID-Net^10^; Free et al. (2025)^11^ |
|  | |  |  |  | | COVID-Net^10^ |
| 18-49 years | | 85.58% | 8.83%  (6.0%, 12.8%)^c^ | 5.60%  (3.9%, 6.6%)^c^ | |  |
| 50-64 years | | 81.25% | 9.28%  (8.6%, 10.7%)^c^ | 9.48%  (6.9%, 12.2%)^c^ | |  |
| 65+ years | | 85.30% | 8.73%  (9.9%, 7.0%)^c^ | 5.98%  (3.6%, 10.3%)^c^ | |  |
| **In-Hospital Mortality** | | | | | | |
| **Age Group** | | **No ICU or MV** | **ICU Only** | **ICU with MV** | |  |
| 6 months – 4 years | | 0.04% | 0.62% | 3.18% | | University of Michigan COVID-19 Vaccination Modeling Team analysis of (March 2022 – October 2023)^13^; Andersen et al. (2025)^15^ |
| 5 -17 years | | 0.10% | 1.46% | 7.57% | | University of Michigan COVID-19 Vaccination Modeling Team analysis of (March 2022 – October 2023)^13^; Andersen et al. (2025)^15^ |
| 18-49 years (high-risk) | | 0.21%  (0.19%, 0.23%) ^d^ | 1.97% (1.77%, 2.17%) ^d^ | 25.37% (22.8%, 27.9%) ^d^ | | University of Michigan COVID-19 Vaccination Modeling Team analysis of (March 2022 – October 2023) ^13^; Joshi et al. (2025) ^14^ |
| 50-64 years (high-risk) | | 0.62%  (0.56%, 0.68%) ^d^ | 4.55% (4.10%, 5.01%) ^d^ | 37.81% (34.0%, 41.6%)^d^ | |  |
| 65+ years (all) | | 1.80% (1.62%, 1.98%) ^d^ | 13.1% (11.8%, 14.4%) ^d^ | 53.3% (48.0%, 58.6%)^d^ | |  |
| **Percentage with Long COVID^†^** | | | | | | |
| 6 months -17 years | 0% | | | | Assumption | |
| 18-49 years | 8.5% (7.0%, 10.2%)^e^ | | | | CDC Post-COVID Conditions^4^ | |
| 50-64 years | 9.6% (8.4%, 11.0%)^e^ | | | |  |  |
| 65+ years | 8.1% (5.9%, 10.9%)^e^ | | | |  |  |

| **Probability of Infection-induced Myocarditis^f^** | | |
| --- | --- | --- |
| 0-17 years | 0.1220% | Boehmer (2021) ^21^ |
| 18-49 years | 0.0793% |  |
| 50-64 years | 0.1370% |  |
| 65+ years | 0.1800% |  |

ICU: intensive care unit; MV; mechanically ventilated; CDC Centers for Disease Control and Prevention; EHR; electronic health record

*General population estimates used for all patients ≥18 years

^†^General population values used for high-risk ages 18-64 years

^a^Varied ±25% of the base-case values

^b^Range for high-risk patients calculated using 95% CI for RR for outpatient care (RR=1.16, RR=1.78)

^c^Range based on minimum and maximum from COVID-NET data

^d^Varied ±10% of the base-case values

^e^Range based on 95% CI

^f^Assumes 50% female for all ages

### 6 Adverse Events

It was assumed that patients receiving no vaccine would not experience adverse events. Age-specific Grade 3 and 4 Local and Systemic adverse event (AE) rates for mRNA-1273 were estimated from Moderna clinical trial data.^22-24^ AE rates for BNT162b2 were assumed to be equivalent to mRNA-1273. All vaccines are also associated with a risk of myocarditis/pericarditis^25^ and anaphylaxis.^26^

Table 13. Vaccine-related adverse event rates for mRNA-1273*

| **Probability** | **6 mos to 4 years** | **5 – 17 years^†^** | **≥18 years** | **Source** |
| --- | --- | --- | --- | --- |
| Grade 3 Local | 0.71% | 3.36% | 4.90% | mRNA-1273-P204 trial data^22^; Berthaud et al. (2024)^23^; Chu et al. (2022)^24^ |
| Grade 4 Local | 0%** | 0%** | 0% |  |
| Grade 3 Systemic | 2.14% | 6.04% | 7.61% |  |
| Grade 4 Systemic | 0%** | 0%** | 0% |  |
| Anaphylaxis | 0% | 0.00049% | 0.00049% | Klein et al. (2021)^26^ |
| Myocarditis/Pericarditis | 0.0008% | 0.0008% | 0.0008% | FDA Letter^25^ |

*AE rates for BNT162b2 (ages 18+) were assumed equal to mRNA_1273

†AE rates for 5-17 years represent a weighted average of the 6-11 year old data presented by Berthaud et al. (2024) and 12-17 year old estimates, which were assumed to be equal to the ≥18 years data from Chu et al. (2022)

**No Grade 4 AEs were assumed for mRNA-1273 in those <18 years of age due to small sample sizes in the mRNA-1273-P204 clinical trial

### 7 Cost Inputs

Table 14. Adverse event costs

| **Adverse Event** | **Cost** | **Source** |
| --- | --- | --- |
| Grade 3 Local* | $6.47 | Berthaud et al. (2024)^23^; Chu et al. (2022)^24^  CMS 2025 Physician’s Fee Schedule^27^  Drugs.com^28^  Rousculp MD, et al. 2024^29^  Walmart.com^30^ |
| Grade 3 Systemic* | $6.47 | Berthaud et al. (2024)^23^; Chu et al. (2022)^24^; CMS 2025 Physician’s Fee Schedule^27^  Drugs.com^28^  Rousculp MD, et al. 2024^29^  Walmart.com^30^ |
| Grade 4 Local | $3,810 | CMS 2025 Physician’s Fee Schedule^27^  CMS Quarterly Addenda Updates^31^  Prosser et al. 2019^32^ |
| Grade 4 Systemic | $3,810 | CMS 2025 Physician’s Fee Schedule^27^  CMS Quarterly Addenda Updates^31^  Prosser et al. 2019^32^ |
| Anaphylaxis | $7,262 | CDC MMWR (2021)^33^  HCUPnet Hospital Inpatient National Statistics^34^  CMS 2025 Physician’s Fee Schedule^27^  CMS Quarterly Addenda Updates^31^  Tutle (2020)^35^ |
| Myocarditis/Pericarditis | $12,120 | HCUPnet Hospital Inpatient National Statistics^36^  HCUPnet Average Hospital Costs per ED Visit^37^  CMS 2025 Physician’s Fee Schedule^27^ |

CDC, Center for Disease Control; CMS, Centers for Medicare and Medicaid Services; MMWR, Mortality and Morbidity weekly report.

*For those ages ≥12 years, based on Chu et al. (2022), it was assumed that 4.6% of grade 3 local and systemic AEs would require an outpatient visit and prescription pain medication for those ages ≥12 years. For those <5 years and 6-11 years, it was estimated that 56.9% and 41.8%^23^ of grade 3 local and systemic AEs would require an outpatient visit, respectively, and over the counter medication only. The cost of an outpatient visit ($54.99) was estimated from 2025 CMS data.^27^ The cost of prescription pain medications (30 oral tablets of 300 mg acetaminophen/30 mg codeine) was $11.60 for those ≥12 years.^28^ Rousculp et al. (2024)^29^ estimated that 22.7% of individuals required over the counter medication use for the 6 days post COVID-19 vaccination. The cost of 6 pills of acetaminophen was estimated at $0.12 for those ≥12 years.^38^ Outpatient medication costs (acetaminophen oral suspension 160 mg per 5mL) were estimated to be $0.88 for those <5 and $1.47 for those 6-11 years^30^; these costs were applied to all AEs requiring an outpatient visit for those <12 years of age.

Table 15. Hospitalization costs by location of care

| **Initial Hospitalization Costs** | **General Population Cost*** | **High-Risk Cost*** | **Cost** | **Source** |
| --- | --- | --- | --- | --- |
| No ICU or Ventilator | $13,351 | $13,585 | $13,537 | Yehoshua A, et al. 2024^39^; Kapinos et al.^40^ |
| ICU only | $22,140 | $22,527 | $22,449 |  |
| ICU with Ventilator | $50,707 | $51,594 | $51,415 |  |

ICU, Intensive care unit.

* General population cost: This cost was used for the ≥65 years population; High-risk cost: General population cost multiplied by 1.07 based on a RR from Kapinos et al.^40^ used for the 12- 64 years high-risk population.

** Final costs: As the model does not allow for age-specific costs, a weighted average was calculated using the population size shown in Table 10.

Table 16. Outpatient and hospital recovery costs

| **Outpatient Costs** | **Average Number of Visits**  **(Per Patient)** | **Cost** | **Source** |
| --- | --- | --- | --- |
| Outpatient visit cost | 1.0 | $191 | Optum Database Analysis (v09062024)^41^ |
| Emergency department visit cost | 1.0 | $495 |  |
| Weighted average | | $685 |  |

Table 17. Post-Infection costs by initial treatment location

| **Treatment location** | **Cost:**  **All patients** | **Cost:**  **High Risk Patients** | **Weighted Average Cost** | **Source** |
| --- | --- | --- | --- | --- |
| Outpatient Care | $843.60 | $2,971.87 | $2,542 | All patients:  Chambers et al. 2023^42^  High Risk patients:  Scott et al. 2024^43^ |
| Hospitalized | $1,085.98 | $16,272.25 | $13,202 |  |

Table 18. Infection-related Myocarditis costs

|  | **Cost** | **Source** |
| --- | --- | --- |
| Infection-related myocarditis | $33,889 | HCUPnet Hospital Inpatient National Statistics. 2018^36^  CMS 2025 Physician’s Fee Schedule^27^ |

### 8 QALY Inputs

Table 19. Adverse event QALY losses

| **Adverse Event** | **QALY loss** | **Source** |
| --- | --- | --- |
| Grade 3 Local | 0.0004 | Walter EB, et al. (2024)^44^ |
| Grade 4 Local | 0.0019 | Assumption based on Prosser et al. (2019)^32*^ |
| Grade 3 Systemic | 0.0004 | Walter EB, et al. (2024)^44^ |
| Grade 4 Systemic | 0.0019 | Assumption based on Prosser et al. (2019)^32*^ |
| Anaphylaxis | 0.0019 | Assumption based on Prosser et al. (2019)^32*^ |
| Myocarditis/Pericarditis^‡^ | 0.0006 | Assumption based on Prosser et al. (2019)^32*‡^ |

LOS, length of stay; QALD, quality-adjusted life days; QALY, quality-adjusted life year.

*0.7 QALD lost due to severe AE of 3 days.

‡Median LOS due to vaccine induced myocarditis/pericarditis is 1 day (0.7 QALD/3).

Table 20. QALY lost due to infection

| **Location of Treatment** | | **QALY loss** | **Source** |
| --- | --- | --- | --- |
| **<18 years of age** | | | |
| No Formal Care | | 0.0102 | University of Michigan COVID-19 Vaccination Modeling Team^13^; Assumption (equal to outpatient care estimate) |
| Outpatient Care | | 0.0102 | Mercon et al., (2025),^45^ University of Michigan COVID-19 Vaccination Modeling Team^13^ |
| Hospitalized: | |  |  |
| No ICU or Ventilator | | 0.0320 | Mercon et al., (2025),^45^ University of Michigan COVID-19 Vaccination Modeling Team^13^ |
| ICU only | | 0.1545 |  |
| Ventilator | | 0.1545 |  |
| Readmission: | |  |  |
| No ICU or Ventilator | | 0.0320 | Assumption (equal to QALY loss of initial hospitalization) |
| ICU only | | 0.1545 |  |
| Ventilator | | 0.1545 |  |
| **18+ years of age** | | | |
| No Formal Care | | 0.0046 | University of Michigan COVID-19 Vaccination Modeling Team^13^; Assumption (equal to outpatient care estimate) |
| Outpatient Care | | 0.0046 | University of Michigan COVID-19 Vaccination Modeling Team^13^ |
| Hospitalized: | |  |  |
| No ICU or Ventilator | | 0.0174 | University of Michigan COVID-19 Vaccination Modeling Team^13^ |
| ICU only | | 0.0394 |  |
| Ventilator | | 0.0394 |  |
| Readmission: | |  |  |
| No ICU or Ventilator | | 0.0174 | Assumption (equal to QALY loss of initial hospitalization) |
| ICU only | | 0.0394 |  |
| Ventilator | | 0.0394 |  |
| **All Ages** | | | |
| No Formal Care | 0 | | Assumption |
| Outpatient Care | 0.0231 | | Sandmann et al. (2022)^46^, adjusted for outpatient care QALY loss |
| Hospitalized | 0.1026 | | PHOSP (2022)^47^ |
| Infection-related myocarditis | 0.0019 | | Assumption based on Prosser et al. (2019)^32*^ |

ICU, intensive care unit; QALY, quality-adjusted life year.

*0.7 quality-adjusted life-days (QALD) lost due to severe adverse event of 3 days.

Table 21. Baseline utility by age group

| **Age group** | **Baseline utility data for population** | **Source** |
| --- | --- | --- |
| 0-4 years | 0.9205 | Hanmer et al. (2006)^48^ |
| 5-17 years | 0.9205 |  |
| 18-49 years | 0.9055 |  |
| 50-64 years | 0.8662 |  |
| 65-100 years | 0.8092 |  |

### 9 Lost Productivity (Patient)

Table 22. Productivity loss

| **Model Parameter** | **Value** | **Data Source** |
| --- | --- | --- |
| Percentage of adults in US labor force |  |  |
| 18-49 years* | 61.5% | US Bureau of Labor Statistics^49^ |
| 50-64 years* | 55.9% | US Bureau of Labor Statistics^49^ |
| ≥65 years | 18.6% | US Bureau of Labor Statistics^49^ |
| Daily wage | $292 | US Bureau of Labor Statistics^50^ |
| Time loss (days) due to: |  |  |
| Vaccination | 0.043 | Prosser et al. (2019)^32^, CDC^51^ |
| Grade 3 Local | 0.19 | Rousculp et al. (2024)^29^ |
| Grade 3 Systemic | 0.19 | Rousculp et al. (2024)^29^ |
| Grade 4 Local** | 1.39 | Prosser et al. (2019)^32^ |
| Grade 4 Systemic** | 1.39 | Prosser et al. (2019)^32^ |
| Anaphylaxis | 2.25 | HCUPnet^34^ |
| Myocarditis/Pericarditis | 1.00 | Shimabukuro (2022)^52^ |
| Infection-related myocarditis | 3.0 | HCUPNet^36^ |
| Short-term infection period: |  |  |
| No formal care | 3.3 | Zheng et al. (2025)^53^ |
| Outpatient care only | 3.3 | Zheng et al. (2025)^53^ |
| Hospitalization: |  |  |
| Symptoms prior to hospitalization | 3.3 | Assumption (equal to outpatient care) |
| In-hospital | 5.71 |  |
| Recovery | 8.8 | Chopra et al. (2021)^54^; Yang et al. (2024)^55^ |
| Readmission | 5.71 | Assumption (equal to original length of stay) |
| Long COVID |  |  |
| Sought outpatient care only for acute infection | 14.58 | Bartsch et al. (2025)^56^ |
| Hospitalized during acute infection | 14.58 | Bartsch et al. (2025)^56^ |

*Based on the Minnesota Department of Health Diabetes unit, 19% of patients with diabetes report being unable to work; this estimate was used to reduce the general population labor force participation rates for those at high-risk (ages 18-49 and 50-64).

**Assumed 50% of patients visit the ER and 50% require hospitalizations.

### 10 Lost Productivity (Caregiver)

Table 23. Caregiver productivity loss

| **Model Parameter** | **Value** | **Data Source** |
| --- | --- | --- |
| Percentage of adults in US labor force | 75.9% | US Bureau of Labor Statistics^49^ |
| Daily wage | $292 | US Bureau of Labor Statistics^50^ |
| Time loss (days) due to: |  |  |
| Vaccination | 0.083 | Prosser et al. (2019)^32^, CDC^51^ |
| Grade 3 Local | 0.19 | Rousculp et al. (2024)^29^ |
| Grade 3 Systemic | 0.19 | Rousculp et al. (2024)^29^ |
| Grade 4 Local** | 1.39 | Prosser et al. (2019)^32^ |
| Grade 4 Systemic** | 1.39 | Prosser et al. (2019)^32^ |
| Anaphylaxis | 2.25 | HCUPnet^34^ |
| Myocarditis/Pericarditis | 1.00 | Shimabukuro (2022)^52^ |
| Infection-related myocarditis | 3.0 | HCUPNet^36^ |
| Short-term infection period: |  |  |
| No formal care | 0.80 | Assumed 50% of outpatient care |
| Outpatient care only | 1.59 | Ortega-Sanchez^57^, Optum database^19^ |
| Hospitalization: |  |  |
| Symptoms prior to hospitalization | 0 | Assumption |
| In-hospital | 7.71 | Hutton (ACIP June 2023)^58^ |
| Recovery | 0 | Assumption |
| Readmission | 7.71 | Assumption (equal to original length of stay) |
| Long COVID |  |  |
| Sought outpatient care only | 14.58 | Bartsch et al. (2025)^56^ |
| Hospitalized | 14.58 | Bartsch et al. (2025)^59^ |

**Assumed 50% of patients visit the ER and 50% require hospitalizations.

### 11 Details on Scenario Analyses

#### 11.1 Inputs for VE Scenario Analyses

Table 24. mRNA-1273 initial VE inputs used for scenario analyses*

| **Scenario** | **0-4 years** | | **5-17 years** | | **18-49 years** | | **50-64 years** | | **65+ years** | |
| --- | --- | --- | --- | --- | --- | --- | --- | --- | --- | --- |
|  | **Infection** | **Hospitalization** | **Infection** | **Hospitalization** | **Infection** | **Hospitalization** | **Infection** | **Hospitalization** | **Infection** | **Hospitalization** |
| ICATT | 84.4% | 84.4% | 62.4% | 62.4% | 57.0% | 50.5% | 57.0% | 50.5% | 57.0% | 57.2% |

*Base-case estimates used for those <18; ICATT values used for those ≥18 years

### 12 Clinical Results for mRNA-1273 compared to BNT162b2

Table 25. Cases Averted (mRNA-1273 relative to BNT162b2)

| **Strategy** | **Symptomatic Infections** | **Outpatient** | **Long COVID** | **Hospitalizations** | **Death** |
| --- | --- | --- | --- | --- | --- |
| 18-64 High-Risk, 65+ All (1-Dose) | 539,125 | 206,693 | 19,740 | 38,476 | 5,067 |
| 18-64 High-Risk (1-Dose), 65+ All (2-Doses) | 544,895 | 210,242 | 20,026 | 38,476 | 5,067 |
| 18-64 High-Risk Subgroup  (1-Dose) | 193,184 | 30,372 | 2,968 | 2,054 | 233 |
| 65+ All Subgroup  (1-Dose) | 345,941 | 176,321 | 16,771 | 36,422 | 4,834 |

### 13 Deterministic Sensitivity Analysis Results

Figure 3. Tornado diagram for clinical outcomes (mRNA-1273 relative to no vaccination)

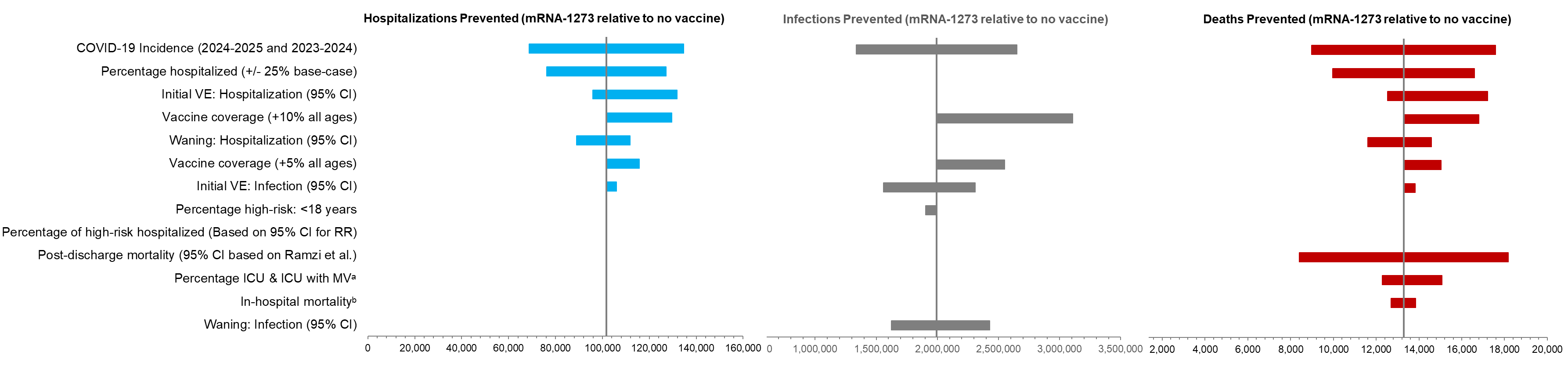
CI: confidence interval; ICU: intensive care unit; RR: relative risk; VE: vaccine efficacy.

a. Minimum and maximum based on COVID-NET data.

b. Lower and upper bounds based on UM-CDC data.

Table 26. Clinical Results of Deterministic Sensitivity Analysis Results (mRNA-1273 relative to no vaccination)

| **Model Parameter** | **Scenario** | **Symptomatic infections averted** | | **Hospitalisations averted** | | **Deaths averted** | |
| --- | --- | --- | --- | --- | --- | --- | --- |
|  |  | **Number** | **% Change from Base** | **Number** | **% Change from Base** | **Number** | **% Change from Base** |
| Base Case | | 1,992,850 | - | 101,739 | - | 13,285 | - |
| COVID-19 Incidence | 2023-2024 Incidence (upper bound) | 2,650,256 | 32.99% | 134,684 | 32.38% | 17,583 | 32.35% |
| Initial VE (Hospitalization) | Upper bound based on 95% CI | 1,992,735 | -0.01% | 131,907 | 29.65% | 17,212 | 29.56% |
| Vaccine coverage | Increase coverage by 10% (all ages) | 3,106,948 | 55.90% | 129,579 | 27.36% | 16,798 | 26.44% |
| Percentage hospitalized | Upper Bounds (+25% of base-case) | 1,992,680 | -0.01% | 127,156 | 24.98% | 16,604 | 24.98% |
| Vaccine coverage | Increase coverage by 5% (all ages) | 2,549,899 | 27.95% | 115,659 | 13.68% | 15,041 | 13.22% |
| Vaccine coverage | Increase coverage by 5% (18+ only) | 2,375,149 | 19.18% | 115,005 | 13.04% | 14,987 | 12.81% |
| Waning (hospitalization) | Lower bound: 1.37% | 1,992,827 | 0.00% | 111,730 | 9.82% | 14,587 | 9.80% |
| Initial VE (infection) | Upper bound based on 95% CI | 2,310,827 | 15.96% | 105,927 | 4.12% | 13,823 | 4.05% |
| Post-discharge mortality | Lower Bound: 2.78% (95% CI lower bound based on Ramzi et al.) | 1,993,105 | 0.01% | 101,759 | 0.02% | 8,400 | -36.77% |
| Percentage ICU & ICU with MV | Lower bounds (minimum based on COVID-NET data) | 1,992,901 | 0.00% | 101,743 | 0.00% | 12,291 | -7.48% |
| In-hospital mortality | Lower bound (based on UM-CDC) | 1,992,882 | 0.00% | 101,741 | 0.00% | 12,691 | -4.47% |
| Initial VE (infection) | Lower bound based on 95% CI | 1,558,190 | -21.81% | 101,739 | 0.00% | 13,285 | 0.00% |
| Waning (Infection) | Lower bound: 3.05% | 2,428,765 | 21.87% | 101,739 | 0.00% | 13,285 | 0.00% |
| Waning (Infection) | Upper bound: 6.75% | 1,623,283 | -18.54% | 101,739 | 0.00% | 13,285 | 0.00% |
| In-hospital mortality | Upper bounds (based on UM-CDC) | 1,992,822 | 0.00% | 101,736 | 0.00% | 13,845 | 4.22% |
| Percentage ICU & ICU with MV | Upper bounds (maximum based on COVID-NET data) | 1,992,762 | 0.00% | 101,731 | -0.01% | 15,074 | 13.47% |
| Percentage of high-risk hospitalized | <18 same as base-case 18+: Upper bound based on 95% CI for RR (RR=1.72)  18-49: 0.33% 50-64: 1.23% ≥65: 8.29.% | 1,992,851 | 0.00% | 101,727 | -0.01% | 13,283 | -0.02% |
| Post-discharge mortality | Upper Bound: 6% (12.96% CI upper bound based on Ramzi et al.) | 1,992,596 | -0.01% | 101,718 | -0.02% | 18,168 | 36.76% |
| Percentage of high-risk hospitalized | <18 same as base-case 18+ Lower bound based on 95% CI for RR (RR=1.66)  18-49: 0.33% 50-64: 1.22% ≥65: 8.29% | 1,992,851 | 0.00% | 101,670 | -0.07% | 13,276 | -0.07% |
| Percentage high-risk (<18) | 28.1% based on Wisk et al. | 1,903,092 | -4.50% | 101,466 | -0.27% | 13,262 | -0.17% |
| Initial VE (Hospitalization) | Lower bound based on 95% CI | 1,992,873 | 0.00% | 95,932 | -5.71% | 12,525 | -5.72% |
| Waning (hospitalization) | Upper bound: 3.87% | 1,992,880 | 0.00% | 88,898 | -12.62% | 11,611 | -12.60% |
| Percentage hospitalized | Lower Bounds (-25% of base-case) | 1,993,021 | 0.01% | 76,315 | -24.99% | 9,965 | -24.99% |
| COVID-19 Incidence | 2024-2025 Incidence (lower bound) | 1,335,252 | -33.00% | 68,775 | -32.40% | 8,985 | -32.37% |

CI: confidence interval; ICU: intensive care unit; MV: mechanical ventilation; RR: relative risk; VE: vaccine efficacy.

Table 27. Economic Results of Deterministic Sensitivity Analysis Results (mRNA-1273 relative to no vaccination)

| **Model Parameter** | **Scenario** | **ICER** | **% Change from Base** |
| --- | --- | --- | --- |
| **Base Case** | | **$23,265** | **-** |
| COVID-19 Incidence | 2024-2025 Incidence (lower bound) | $63,587 | 173.31% |
| Percentage hospitalized | Lower Bounds (-25% of base-case) | $40,177 | 72.69% |
| Post-discharge mortality | Lower Bound: 2.78% (95% CI lower bound based on Ramzi et al.) | $39,272 | 68.80% |
| Vaccine administration cost | $40 based on CMS vaccine pricing payment allowance | $32,902 | 41.42% |
| Waning (hospitalization) | Upper bound: 3.87% | $31,022 | 33.34% |
| Waning (Infection) | Upper bound: 6.75% | $28,798 | 23.78% |
| Initial VE (infection) | Lower bound based on 95% CI | $28,393 | 22.04% |
| Post-Infection Costs | Lower bound (-25% of base-case) | $28,324 | 21.74% |
| Hospitalization Costs | Lower bound (-25% of base-case) | $26,538 | 14.07% |
| Initial VE (Hospitalization) | Lower bound based on 95% CI | $26,437 | 13.63% |
| Percentage ICU & ICU with MV | Lower bounds (minimum based on COVID-NET data) | $26,093 | 12.16% |
| Percentage no formal care | Upper Bounds (calculated using RR=1.16 for high-risk outpatient care; ≥65 years +25% of base-case) | $25,644 | 10.22% |
| In-hospital mortality | Lower bound (based on UM-CDC) | $24,907 | 7.06% |
| Acute Phase QALYs Lost | Lower bound based on lower bound of ranges from UM-CDC and Mercon (2025) | $24,660 | 6.00% |
| Post-infection QALY loss | Lower bounds (outpatient based on 95% CI; hospitalization -25% of base-case value) | $24,571 | 5.61% |
| Premature mortality productivity losses | Lower bound (-10% of base-case) | $24,517 | 5.38% |
| Vaccine coverage | Increase coverage by 10% (all ages) | $24,376 | 4.78% |
| Outpatient Care Cost | Lower Bound (Equal to UM-CDC values) of $476.92 | $24,185 | 3.96% |
| No Formal Care QALY Loss | Assume 50% of outpatient values | $23,939 | 2.90% |
| Vaccine coverage | Increase coverage by 5% (all ages) | $23,903 | 2.74% |
| Vaccine coverage | Increase coverage by 5% (18+ only) | $23,818 | 2.38% |
| Long COVID Time Loss | Lower bound of 8.11 days from Bartsch | $23,495 | 0.99% |
| Caregiver QALY Loss (<18) | Excluded from acute phase QALY loss estimates for those <18 years | $23,460 | 0.84% |
| Percentage with Long COVID | Lower bounds (95% CI lower bounds) | $23,399 | 0.58% |
| Hospital readmission | Lower bounds - calculated values | $23,334 | 0.30% |
| Percentage of high-risk hospitalized | <18 same as base-case 18+ Lower bound based on 95% CI for RR (RR=1.66)  18-49: 0.33% 50-64: 1.22% ≥65: 8.29% | $23,307 | 0.18% |
| Percentage of high-risk hospitalized | <18 same as base-case 18+: Upper bound based on 95% CI for RR (RR=1.72)  18-49: 0.33% 50-64: 1.23% ≥65: 8.29.% | $23,241 | -0.11% |
| Hospital readmission | Upper bounds - calculated values | $23,224 | -0.18% |
| Percentage with Long COVID | Upper bounds (95% CI upper bounds) | $23,097 | -0.72% |
| Long COVID Time Loss | Upper bound of 21.05 days from Bartsch | $23,035 | -0.99% |
| Hospital Recovery Time Loss | 19.3 days based on Chopra et al. | $22,761 | -2.17% |
| Percentage high-risk (<18) | 28.1% based on Wisk et al. | $22,110 | -4.97% |
| Premature mortality productivity losses | Upper bound (+10% of base-case) | $22,014 | -5.38% |
| Post-infection QALY loss | Upper bounds (outpatient based on 95% CI; hospitalization +25% of base-case value) | $22,012 | -5.39% |
| Acute Phase QALYs Lost | Upper bound based on upper bounds of ranges from UM-CDC and Mercon (2025) | $22,005 | -5.42% |
| In-hospital mortality | Upper bounds (based on UM-CDC) | $21,890 | -5.91% |
| Percentage no formal care | Lower Bounds (calculated using RR=1.78 for high-risk outpatient care; ≥65 years -25% of base-case) | $20,473 | -12.00% |
| Hospitalization Costs | Upper bound (+25% of base-case) | $19,993 | -14.07% |
| Percentage ICU & ICU with MV | Upper bounds (maximum based on COVID-NET data) | $19,308 | -17.01% |
| Waning (hospitalization) | Lower bound: 1.37% | $18,276 | -21.44% |
| Post-Infection Costs | Upper bound (+25% of base-case) | $18,207 | -21.74% |
| Waning (Infection) | Lower bound: 3.05% | $17,328 | -25.52% |
| Initial VE (infection) | Upper bound based on 95% CI | $17,075 | -26.61% |
| Post-discharge mortality | Upper Bound: 6% (12.96% CI upper bound based on Ramzi et al.) | $14,352 | -38.31% |
| Percentage hospitalized | Upper Bounds (+25% of base-case) | $12,139 | -47.82% |
| Initial VE (Hospitalization) | Upper bound based on 95% CI | $9,997 | -57.03% |
| COVID-19 Incidence | 2023-2024 Incidence (upper bound) | $3,148 | -86.47% |

CI: confidence interval; ICU: intensive care unit; MV: mechanical ventilation; QALY: quality-adjusted life year; RR: relative risk; VE: vaccine efficacy.

### 14 Scenario Analysis Results

#### 14.1 mRNA-1273 relative to no vaccination

Table 28. Clinical Scenario Analysis Results (mRNA-1273 relative to no vaccination)

| **Model Parameter** | **Range/Data Selections** | **Symptomatic Infections Averted** | | **Hospitalizations Averted** | | **Deaths Averted** | |
| --- | --- | --- | --- | --- | --- | --- | --- |
|  |  | **Number** | **% Change*** | **Number** | **% Change*** | **Number** | **% Change*** |
| Base-Case | | 1,992,850 | - | 101,739 | - | 13,285 | - |
| VE | VE against symptomatic COVID-19 infection for adults ages 18 years and older from ICATT study | 2,342,539 | 17.55% | 103,366 | 1.60% | 13,471 | 1.40% |
| Percentage hospitalized (high-risk) | Increased RR of hospitalization (RR=1.45) for high-risk (applies to 18+ only) | 1,992,852 | 0.00% | 101,530 | -0.21% | 13,262 | -0.17% |
| Inpatient mortality (high-risk) | Increased mortality (RR=1.62) for high-risk (18+ only) | 1,992,853 | 0.00% | 101,739 | 0.00% | 13,255 | -0.23% |
| Vaccine coverage rates | Increase coverage by 5% for all ages | 2,549,899 | 27.95% | 115,659 | 13.68% | 15,041 | 13.22% |
| Vaccine coverage rates | Increase coverage by 10% for all ages | 3,106,948 | 55.90% | 129,579 | 27.36% | 16,798 | 26.44% |
| Vaccine coverage rates | Increase coverage by 5% for 18+ only | 2,375,149 | 19.18% | 115,005 | 13.04% | 14,987 | 12.81% |
| Indirect benefit | Based on CDC Right Study | 2,131,029 | 6.93% | 101,739 | 0.00% | 13,285 | 0.00% |

CDC: Centers for Disease Control; RR: relative risk

*From Base-Case Results

#### 14.2 mRNA-1273 relative to BNT162b2

Table 29. Clinical Scenario Analysis Results (mRNA-1273 relative to BNT162b2)

| **Model Parameter** | **Range/Data Selections** | **Symptomatic Infections Averted** | | **Hospitalizations Averted** | | **Deaths Averted** | |
| --- | --- | --- | --- | --- | --- | --- | --- |
|  |  | **Number** | **% Change*** | **Number** | **% Change*** | **Number** | **% Change*** |
| Strategy: 18-64 High-Risk, 65+ All (1-Dose) | | | | | | | |
| Base-Case | | 539,125 | - | 38,476 | - | 5,067 | - |
| COVID-19 Incidence | 2023-2024 Incidence (upper bound) | 714,762 | 32.6% | 51,166 | 33.0% | 6,737 | 33.0% |
| COVID-19 Incidence | 2024-2025 Incidence (lower bound) | 363,415 | -32.6% | 25,779 | -33.0% | 3,395 | -33.0% |
| Initial VE (infection) | Upper bound for BNT162b2 based on 95% CI  18-64: 47.7%  65+: 48.3% | 159,370 | -70.4% | 18,019 | -53.2% | 2,379 | -53.1% |
| Initial VE (infection) | Lower bound for BNT162b2 based on 95% CI  18-64: 35.5%  65+: 26.6% | 932,505 | 73.0% | 40,260 | 4.6% | 5,287 | 4.3% |
| Initial VE (Hospitalization) | Upper bound for BNT162b2 based on 95% CI  18-64: 45.7%  65+: 52.0% | 539,243 | 0.0% | 11,353 | -70.5% | 1,485 | -70.7% |
| Initial VE (Hospitalization) | Lower bound for BNT162b2 based on 95% CI  18-64: 30.3%  65+: 19.3% | 539,125 | 0.0% | 38,476 | 0.0% | 5,067 | 0.0% |
| Vaccine coverage | Increase coverage by 5% (all ages) | 638,384 | 18.4% | 43,435 | 12.9% | 5,712 | 12.7% |
| Vaccine coverage | Increase coverage by 10% (all ages) | 737,642 | 36.8% | 48,395 | 25.8% | 6,357 | 25.5% |
| Strategy: 65+ All Subgroup (1-Dose) | | | | | | | |
| Base-Case | | 345,941 | - | 36,422 | - | 4,834 | - |
| COVID-19 Incidence | 2023-2024 Incidence (upper bound) | 455,169 | 31.6% | 48,392 | 32.9% | 6,422 | 32.9% |
| COVID-19 Incidence | 2024-2025 Incidence (lower bound) | 236,645 | -31.6% | 24,444 | -32.9% | 3,244 | -32.9% |
| Initial VE (infection) | Upper bound for BNT162b2 based on 95% CI  18-64: 47.7%  65+: 48.3% | 142,479 | -58.8% | 17,354 | -52.4% | 2,303 | -52.4% |
| Initial VE (infection) | Lower bound for BNT162b2 based on 95% CI  18-64: 35.5%  65+: 26.6% | 560,214 | 61.9% | 37,353 | 2.6% | 4,957 | 2.6% |
| Initial VE (Hospitalization) | Upper bound for BNT162b2 based on 95% CI  18-64: 45.7%  65+: 52.0% | 346,054 | 0.0% | 10,209 | -72.0% | 1,355 | -72.0% |
| Initial VE (Hospitalization) | Lower bound for BNT162b2 based on 95% CI  18-64: 30.3%  65+: 19.3% | 345,941 | 0.0% | 36,422 | 0.0% | 4,834 | 0.0% |
| Vaccine coverage | Increase coverage by 5% (all ages) | 385,228 | 11.4% | 40,797 | 12.0% | 5,414 | 12.0% |
| Vaccine coverage | Increase coverage by 10% (all ages) | 475,640 | 37.5% | 51,463 | 41.3% | 6,830 | 41.3% |

CI: Confidence interval; VE: Vaccine efficacy.

Table 30. Economic Scenario Analysis Results (mRNA-1273 relative to BNT162b2)

| **Model Parameter** | **Range/Data Selections** | **ICER** | **% Change*** |
| --- | --- | --- | --- |
| Strategy: 18-64 High-Risk, 65+ All (1-Dose) | | | |
| Base-Case | | 1273 dominates BNT162b2 | - |
| COVID-19 Incidence | 2023-2024 Incidence (upper bound) | 1273 dominates BNT162b2 | N/A |
| COVID-19 Incidence | 2024-2025 Incidence (lower bound) | 1273 dominates BNT162b2 | N/A |
| Initial VE (infection) | Upper bound for BNT162b2 based on 95% CI  18-64: 47.7%  65+: 48.3% | 1273 dominates BNT162b2 | N/A |
| Initial VE (infection) | Lower bound for BNT162b2 based on 95% CI  18-64: 35.5%  65+: 26.6% | 1273 dominates BNT162b2 | N/A |
| Initial VE (Hospitalization) | Upper bound for BNT162b2 based on 95% CI  18-64: 45.7%  65+: 52.0% | 1273 dominates BNT162b2 | N/A |
| Initial VE (Hospitalization) | Lower bound for BNT162b2 based on 95% CI  18-64: 30.3%  65+: 19.3% | 1273 dominates BNT162b2 | N/A |
| Vaccine coverage | Increase coverage by 5% (all ages) | 1273 dominates BNT162b2 | N/A |
| Vaccine coverage | Increase coverage by 10% (all ages) | 1273 dominates BNT162b2 | N/A |
| Strategy: 65+ All Subgroup (1-Dose) | | | |
| Base-Case | | 1273 dominates BNT162b2 | - |
| COVID-19 Incidence | 2023-2024 Incidence (upper bound) | 1273 dominates BNT162b2 | N/A |
| COVID-19 Incidence | 2024-2025 Incidence (lower bound) | 1273 dominates BNT162b2 | N/A |
| Initial VE (infection) | Upper bound for BNT162b2 based on 95% CI  18-64: 47.7%  65+: 48.3% | 1273 dominates BNT162b2 | N/A |
| Initial VE (infection) | Lower bound for BNT162b2 based on 95% CI  18-64: 35.5%  65+: 26.6% | 1273 dominates BNT162b2 | N/A |
| Initial VE (Hospitalization) | Upper bound for BNT162b2 based on 95% CI  18-64: 45.7%  65+: 52.0% | 1273 dominates BNT162b2 | N/A |
| Initial VE (Hospitalization) | Lower bound for BNT162b2 based on 95% CI  18-64: 30.3%  65+: 19.3% | 1273 dominates BNT162b2 | N/A |
| Vaccine coverage | Increase coverage by 5% (all ages) | 1273 dominates BNT162b2 | N/A |
| Vaccine coverage | Increase coverage by 10% (all ages) | 1273 dominates BNT162b2 | N/A |

CI: Confidence interval; N/A: Not Applicable; VE: Vaccine efficacy.

### 16 Antimicrobial Resistance

Antimicrobial resistance (AMR) is a global health threat where microorganisms, such as bacteria, viruses, fungi, and parasites, become resistant to the drugs designed to treat them. Antibiotics can save lives, but any time they are used they can contribute to resistance.

Antibiotic prescriptions received in the outpatient setting for COVID-19 patients has been reported in multiple studies.^60,61^ Table 33 displays the proportion of COVID-19 patients in the outpatient setting estimated to receive antibiotic prescriptions.

Table 31. Proportion of COVID-19 outpatients receiving antibiotic prescriptions

| **Age group** | **Proportion prescribed antibiotics** | **Source** |
| --- | --- | --- |
| 0-4 years | 3.60% | Extrapolated from Figure 2 in Wittman et al. 2023^60^ |
| 5-17 years | 5.27% | Extrapolated from Figure 2 in Wittman et al. 2023^60^ for age groups 0-5 and 6-17, and then weighted by US population size. |
| 18-49 years | 11.51% | Extrapolated from Figure 2 in Wittman et al. 2023^60^ for age groups 18-24, 25-44, and 45-64, and then weighted by US population size. |
| 50-64 years | 16.00% | Extrapolated from Figure 2 in Wittman et al. 2023^60^ |
| 65+ years | 29.90% | Based on data from Tsay et al. 2022^61^. Weighted average based on US population size for age groups. |

The number of antibiotic prescriptions that could be avoided with use of mRNA-1273 compared to no vaccine is shown in Table 34.

Table 32. Number of antibiotic prescriptions avoided

| **Age groups** | **Number of Prescriptions** | | **Prescriptions avoided** |
| --- | --- | --- | --- |
|  | **No Vaccine** | **1273** |  |
| 0-4 years | 8,597 | 8,347 | 250 |
| 5-17 years | 5,579 | 5,284 | 295 |
| 18-49 years | 181,721 | 175,808 | 5,913 |
| 50-64 years | 261,821 | 246,068 | 15,753 |
| 65-100 years | 898,344 | 775,536 | 122,808 |
| Total | 1,356,062 | 1,211,042 | 145,020 |

### 17 Number Needed to Vaccinate

Table 33. Number needed to vaccinate (mRNA-1273 relative to no vaccination)

| Strategy | Symptomatic Infections | Outpatient | Long COVID | Hospitalizations | Deaths |
| --- | --- | --- | --- | --- | --- |
| 6 months-64 High-Risk, 65+ All (1-Dose) | 31 | 109 | 1,158 | 616 | 4,717 |
| 6 months-4 High-Risk Subgroup (1-Dose) | 4 | 69 | - | 862 | 10,462 |
| 5-64 High-Risk Subgroup (1-Dose) | 29 | 194 | 2,070 | 3,134 | 27,723 |
| 65+ All Subgroup (1-Dose) | 39 | 78 | 809 | 349 | 2,632 |

Table 34. Second Dose: Number needed to vaccinate (mRNA-1273 relative to no vaccination)

| Strategy | Symptomatic Infections | Outpatient | Long COVID | Hospitalizations | Deaths |
| --- | --- | --- | --- | --- | --- |
| 6 months-64 High-Risk, 65+ All (1-Dose): Cases averted | 1,992,850 | 573,144 | 54,141 | 101,739 | 13,285 |
| 6 months-64 High-Risk, 65+ All (2-Dose): Cases averted | 2,030,788 | 594,842 | 56,005 | 103,371 | 13,501 |
| Additional cases averted with 2nd dose | 37,938 | 21,698 | 1,865 | 1,632 | 217 |
| Number of 2nd dose vaccinations | 5,388,691 | | | | |
| NNV with second dose* | 142 | 248 | 2,890 | 3,301 | 24,875 |

*To avert one additional outcome compared to 1-dose strategy.

### 18 Benefit Cost Ratios

Table 35. Benefit Cost Ratios from the Societal Perspective (mRNA-1273 relative to no vaccination)

| **Strategy** | **QALYs Monetized (Value=$100k)** | **QALYs**  **Monetized (Value=$150k)** | **LYs**  **Monetized (VSLY $604k)** | **QALYs**  **Monetized (VQALY $717k)** |
| --- | --- | --- | --- | --- |
| 6 months-64 High-Risk, 65+ All (1-Dose) | 1.91 | 2.48 | 7.60 | 7.90 |
| 6 months-64 High-Risk (1-Dose), 65+ All (2-Doses) | 1.79 | 2.33 | 7.11 | 7.41 |
| 6 months-4 High-Risk Subgroup (1-Dose) | 6.03 | 8.07 | 14.36 | 30.07 |
| 5-64 High-Risk Subgroup (1-Dose) | 0.99 | 1.24 | 3.08 | 3.63 |
| 65+ All Subgroup (1-Dose) | 2.76 | 3.62 | 11.93 | 11.81 |

LY: life year; QALYs: quality-adjusted life years; VSLY: Value of a Statistical Life Year; VQALY: Value per Quality-Adjusted Life Year.

### 20 Cheers Checklist

|  | **Item** | **Guidance for Reporting** | **Reported in section** |
| --- | --- | --- | --- |
| **TITLE** | | |  |
| Title | 1 | Identify the study as an economic evaluation and specify the interventions being compared. | Title Page |
| **ABSTRACT** | | |  |
| Abstract | 2 | Provide a structured summary that highlights context, key methods, results and alternative analyses. | Abstract |
| **INTRODUCTION** | | |  |
| Background and objectives | 3 | Give the context for the study, the study question and its practical relevance for decision making in policy or practice. | Introduction |
| **METHODS** | | |  |
| Health economic  analysis plan | 4 | Indicate whether a health economic analysis plan was developed and  where available. | Analysis plan not developed |
| Study population | 5 | Describe characteristics of the study population (such as age range, demographics, socioeconomic, or clinical characteristics). | Introduction; Methods Overview |
| Setting and location | 6 | Provide relevant contextual information that may influence findings. | Overview |
| Comparators | 7 | Describe the interventions or strategies being compared and why chosen. | Overview |
| Perspective | 8 | State the perspective(s) adopted by the study and why chosen. | Overview |
| Time horizon | 9 | State the time horizon for the study and why appropriate. | Overview |
| Discount rate | 10 | Report the discount rate(s) and reason chosen. | Quality of Life |
| Selection of outcomes | 11 | Describe what outcomes were used as the measure(s) of benefit(s) and harm(s). | Overview; Model Structure |
| Measurement of outcomes | 12 | Describe how outcomes used to capture benefit(s) and harm(s) were measured. | Model Structure and Inputs; Vaccine Effectiveness; |
| Valuation of outcomes | 13 | Describe the population and methods used to measure and value outcomes. | Target Population |
| Measurement and valuation of resources  and costs | 14 | Describe how costs were valued. | Vaccine Unit Costs; Model Structure and Inputs |
| Currency, price date, and conversion | 15 | Report the dates of the estimated resource quantities and unit costs, plus the currency and year of conversion. | Model Structure and Inputs |
| Rationale and  description of model | 16 | If modelling is used, describe in detail and why used. Report if the model  is publicly available and where it can be accessed. | Overview; Model Structure |
| Analytics and assumptions | 17 | Describe any methods for analysing or statistically transforming data, any extrapolation methods, and approaches for validating any model used. | Methods |
| Characterizing heterogeneity | 18 | Describe any methods used for estimating how the results of the study vary for sub-groups. | Methods Overview |
| Characterizing  distributional effects | 19 | Describe how impacts are distributed across different individuals  or adjustments made to reflect priority populations. | Not applicable |
| Characterizing uncertainty | 20 | Describe methods to characterize any sources of uncertainty in the analysis. | Sensitivity Analyses; Scenario Analyses |
| Approach to engagement with patients and others affected by the study | 21 | Describe any approaches to engage patients or service recipients, the general public, communities, or stakeholders (e.g., clinicians or payers) in the design of the study. | Not applicable |
| **RESULTS** | | |  |
| Study parameters | 22 | Report all analytic inputs (e.g., values, ranges, references) including uncertainty or distributional assumptions. | Included in Methods/Tech appendix & DSA/Scenario Results |
| Summary of main results | 23 | Report the mean values for the main categories of costs and outcomes of interest and summarise them in the most appropriate overall measure. | Results |
| Effect of uncertainty | 24 | Describe how uncertainty about analytic judgments, inputs, or projections  affect findings. Report the effect of choice of discount rate and time horizon, if applicable. | Deterministic Sensitivity Analyses; Scenario Analyses |
| Effect of engagement with patients and others affected by the study | 25 | Report on any difference patient/service recipient, general public, community, or stakeholder involvement made to the approach or findings of the study | Not Applicable |
| **DISCUSSION** | | |  |
| Study findings, limitations, generalizability, and current knowledge | 26 | Report key findings, limitations, ethical or equity considerations not captured, and how these could impact patients, policy, or practice. | Discussion |
| **OTHER RELEVANT INFORMATION** | | | |
| Source of funding | 27 | Describe how the study was funded and any role of the funder in the identification, design, conduct, and reporting of the analysis | Transparency |
| Conflicts of interest | 28 | Report authors conflicts of interest according to journal or  International Committee of Medical Journal Editors requirements. | Transparency |

19.	Moderna data on file 2024. Optum’s de-identified Clinformatics® Data Mart Database. Analysis September 2023 to February 2024 COVID-19 medical attendances and hospitalizations.

20.	Moderna. COVID-19 Hospitalization Risk by Comorbidities Profile (Risk Stacking). Moderna bench to practice website. <https://dev.atlas.modernatx.com/bench2practice/Interactive-dashboard>. Accessed.

21.	Boehmer TK, Kompaniyets L, Lavery AM, et al. Association Between COVID-19 and Myocarditis Using Hospital-Based Administrative Data - United States, March 2020-January 2021. *MMWR Morbidity and mortality weekly report.* 2021;70(35):1228-1232.

22.	ModernaTx Inc. *mRNA-1273-P204 Clinical Trial Data.*

23.	Berthaud V, Creech CB, Rostad CA, et al. Safety and Immunogenicity of an mRNA-1273 Booster in Children. *Clinical Infectious Diseases.* 2024;79(6):1524-1532.

24.	Chu L, Vrbicky K, Montefiori D, et al. Immune response to SARS-CoV-2 after a booster of mRNA-1273: an open-label phase 2 trial. *Nature medicine.* 2022;28(5):1042-1049.

25.	FDA U.S. Food & Drug Administration. Letters for myocarditis.

41.	Moderna. Optum Database Analyses (v09062024). Data on File.

1. The COVID-NET data was downloaded on September 12, 2025.
